# Using iterative agentic retrieval-augmented generation to refine the risk of infection from social contexts during the COVID-19 pandemic: Development and Evaluation Study

**DOI:** 10.64898/2026.10.28.26366693

**Authors:** Jonathan Thibaut, Mert Tunç, Johannes De Smedt, Joren Raymenants, Caspar Geenen, Lisa Cavillot, Emmanuel André

**Affiliations:** KU Leuven, Department of Microbiology, Immunology and Transplantation, Laboratory of Clinical Microbiology, Herestraat 49, Leuven 3000, Belgium; KU Leuven, Research Center for Information Systems Engineering (LIRIS), Naamsestraat 69, 3000 Leuven, Belgium; UAntwerpen, Faculteit Geneeskunde en Gezondheidswetenschappen, Drie Eikenstraat 663, 2650 Edegem, Belgium; UZ Leuven - University Hospitals Leuven, Department of Laboratory Medicine, National Reference Centre for Respiratory Pathogens, Herestraat 49, Leuven 3000, Belgium

**Author notes:** **Corresponding Author** Jonathan Thibaut, Laboratory of Clinical Microbiology, KU Leuven, Herestraat 49, 3000 Leuven, Belgium.

**Keywords:** Large Language Models, Contact tracing, COVID-19, Supervised learning, Risk prediction, Conversational agent

## Abstract

**Background:** Contact tracing is crucial to limit pathogen transmission and to gather epidemiological data. However, despite machine learning optimizations, it remains time-consuming. Large language models have been used for infection risk prediction, but no existing application converses with a user to adapt its risk estimates.

**Objective:** We aimed to develop an iterative retrieval-augmented generation conversational framework to predict coronavirus disease 2019 (COVID-19) infection risk from textual social data. The conversational agent aims to gather personalized risk factors based on descriptions of social interactions.

**Methods:** We trained a long short-term memory (LSTM) model to predict the risk of COVID-19 infection from 19,434 textual descriptions of social interactions, collected during the pandemic in the Leuven student community, in Belgium. We integrated this model into a Llama3.3-based conversational framework that iteratively gathers additional social information that may represent a risk of infection. We evaluated the framework using large language model (LLM)-as-a-judge metrics and by comparing its risk assessment in different social contexts.

**Results:** Based solely on social interaction data, a fine-tuned *RobBERT* model predicted infection risk with an area under the receiver operating characteristic curve (ROC AUC) of 0.73. It performed similarly to an LSTM model and a logistic regression on term frequency–inverse document frequency (TF-IDF) features, which both achieved a ROC AUC of 0.72. These three models slightly outperformed a Quantized Low-Rank Adaptation (QLoRA)-fine-tuned *Mistral NeMo* model (ROC AUC = 0.69). *Llama3*.*3* performed well in our framework: the predicted infection risk converged often, and it asked fewer redundant questions than the smaller models (*Mistral 7B, Gemma3 12B*). In addition, the framework showed the ability to stratify, by risk of infection, synthetic users who had different types of social interactions.

**Conclusions:** Combining simple language models with large language models can refine individual infection risk from contact tracing data in a high-circulation context. Our conversational framework may help optimize contact tracing by highlighting social contexts with a high risk of infection.

## Introduction

Contact tracing plays a crucial role in mitigating airborne disease outbreaks. While it helps prevent further transmission by identifying contacts of infected individuals and providing quarantine and isolation advice, it also allows essential epidemiological data gathering to monitor the evolution of the disease [1, 2]. However, contact tracing is time-consuming, expensive, and requires a large workforce. This procedure scales poorly when the number of infected persons drastically increases in the population [3]. This procedure could be optimized with an infection-risk prediction model to prioritize, for instance, the higher-risk contacts of an infected person. Different studies proposed automated contact tracing systems embedding a risk prediction model [4, 5].

In their review, Hang and colleagues highlighted promising results of machine learning-enhanced digital contact tracing to construct contact graphs, to estimate the source of infection, and to predict the risk of infection [4]. Notably, for this latter purpose, Bengio and colleagues used neural networks with set transformer blocks to process symptoms and exposure data recorded over several days. Their simulations suggest that digital contact tracing enhanced with those methods can help decrease the reproduction number [5]. Another approach to estimate the risk of infection relies on a network-based heuristic in contact graphs. Loh and colleagues proposed an application that relies on Wi-Fi, Bluetooth, and ultrasound to notify users about their distance to a positive case in the contact network [6]. Risk prediction can also be enhanced with Deep Graph Infomax, a learning technique to embed vertex and mutual information between vertices in such a graph [7]. Bipartite graphs can help investigate individuals visiting the same venue and their risk of infection. For instance, a simple implementation of the PageRank algorithm could give a high risk score to individuals who visited venues with many positive cases [8]. Also, conversational frameworks which do not rely on machine learning can also improve the contact tracing procedure. Notably, Johnson and colleagues integrated a rule-based conversational software, i.e., not based on LLMs, into a contact tracing system to triage cases. The use of this software reduced the volume of cases requiring a live telephone interview [9]. In a related but public-facing approach, Castonguay and colleagues implemented the Centers for Disease Control and Prevention (CDC) COVID-19 Calculator, a rule-based conversational software that delivered personalized isolation, testing, and masking guidance to people with COVID-19 and their close contacts through a short conversational exchange, reaching millions of unique users [10].

Moreover, the risk of infection may be predicted from the contact tracing notes using natural language processing techniques. Indeed, recent advances in this field have allowed for the development of retrieval-augmented generation (RAG) techniques [11, 12]. A RAG framework combines a generative language model with an external retrieval component. Given an input query, it dynamically fetches relevant documents from a large knowledge source and conditions generation on this evidence to produce specific outputs for a conversation with a user. Typically, the corpus is pre-embedded into vectors and searched by similarity [12]. Such methods have demonstrated the ability to solve complex problems in several medical fields using various foundation models in different natural languages. Notably, in Hong Kong, Yang and colleagues have developed a RAG-based real-time decision support in the context of the COVID-19 outbreak. They used LLMs to extract structured data from mixed databases of structured and unstructured data (e.g., textual epidemiological reports). The data retrieved by the LLMs were used as an input to a reinforcement learning model to better allocate resources [13]. Similarly, LLMs have been studied for their ability to detect outbreaks. For instance, Kaur and Butt proposed an LLM-enhanced framework to detect outbreaks, inform public health decisions, and optimize resource allocations in emergency departments (EDs) [14]. LLMs were also used to assess the risk of a regional outbreak of conjunctivitis based on tweets. Generative Pre-trained Transformer (GPT) models’ assessments achieved a correlation coefficient above 0.60 with experts’ assessments [15]. Also, LLMs have demonstrated promising performance in predicting diagnoses and individual risk of developing a non-communicable disease (NCD), notably by using multi-modal architectures to combine structured and unstructured health data [16]. Niset and colleagues developed a RAG pipeline to support general (communicable and non-communicable diseases) diagnostic patients in the ED. They showed that GPT-4, QWEN2, and Llama3 often agree on the diagnosis. Also, specific diagnoses made by LLMs often matched the specific diagnoses of medical experts [17]. RAG-based risk prediction methods have also been applied specifically for communicable diseases [18]. For instance, Roshani and colleagues showed that a pre-trained LLMs can predict the severity of COVID-19 with an AUC of 0.75 without fine-tuning the model. In their experiments, they serialized structured data as an input to the LLM. They interpreted the predictions of the model by extracting attention scores from the model’s output layer [19].

Šimunović and colleagues showed that gradient boosting and LLMs achieve performance similar to or better than pediatricians in diagnosing serious bacterial infections [20].

While iterative data refinement has been implemented for RAG applications in a general domain, such techniques have not been considered yet for medical applications [21]. In addition, the lack of explainability remains a limitation in the design of conversational agents [16]. Finally, while LLMs have been used in public health and infectious disease modeling [18], to our knowledge, no study has used LLMs to predict infection risk from textual contact-tracing data.

Our study aims to design a conversational framework that iteratively predicts the risk of infection based on textual descriptions of social interactions. The conversational agent must ask relevant questions about the past conversation with the user to refine the infection risk. Also, we aim to keep the system as interpretable as possible.

## 2 Methods

Our conversational risk prediction framework relies on contact tracing data and a supervised learning model. First, we describe the data collection process and the different predictive models evaluated. Second, we describe the architecture of the conversational framework. Third, we present the experimental setup for optimizing the whole conversational architecture. Fourth, we describe the experimental setup for evaluating the performance of the conversational framework in discriminating simulated users by their risk of infection.

### 2.1 Data collection and risk prediction models

During the COVID-19 pandemic, we diagnosed infections with polymerase chain reaction (PCR) tests and performed bidirectional contact tracing in the student population in Leuven, Belgium [22]. The Ethics Committee Research UZ/KU Leuven approved the contact tracing study protocol (approval number S64919). Informed consent was waived, as data collection was limited to public health purposes. Along with this test-and-trace procedure, we collected textual descriptions of social interactions happening before sampling. Those descriptions were either entered by individuals while making an appointment at the test center, or by contact tracers during the phone call procedure. In total, we gathered 30, 312 social event descriptions, linked to at least one PCR diagnostic, which took place between October 5, 2020 and May 19, 2022. Among those, we retained 19, 434 social interactions described by contact tracers and linked to at least one participant with a positive PCR diagnostic (Supplementary Figure 1).

We used this data to train supervised learning models to predict the risk of infection in various social contexts. We studied the performance of LSTM neural networks [23, 24], logistic regression [25], and gradient boosting techniques [26]. Those last two models were trained on data sets preprocessed with the *Word2Vec* and the TF-IDF algorithms [27, 28, 29]. We considered that an event presented a risk of transmission if at least one contact of the positive index case tested positive within 14 days after the social interaction. We integrated the LSTM model within the architecture of the conversational framework. We also conducted experiments to evaluate the performance of more recent transformer-based neural network architectures to assess whether those outperform the more traditional algorithms described earlier. We fine-tuned a pre-trained Dutch RoBERTa-based language model [30] and applied Low-Rank Adaptation (LoRA) to the attention layers of *Mistral NeMo* [31, 32, 33] on our labeled set of textual descriptions of social interactions. We trained both to predict the risk of infection from these data.

### 2.2 Architecture of the conversational framework

The conversational system aims to efficiently estimate the risk of infection. So, it aims to iteratively ask a few relevant questions. Those must be specific to the initial description of the social interactions and must represent potential social risk factors. We implemented a hybrid RAG architecture with sequential steps and autonomous components involving two distinct agents. These agentic components rely on open-weight foundation models, operationalized using *Ollama*, to ensure the users’ privacy. They are essential to assess and summarize information gathered during the conversation with the user. In the experiments presented below, natural language was embedded using *Gemma (7B)* [34], which yielded the best framework performance in our optimization experiments, although *Qwen3 Embedding (8B)* would have been a suitable choice as well (Section 3.2). The behavior of the LLM-based contact tracing system was implemented using *LangChain* and *LangGraph1*.*0*.*9 Python3*.*10* packages. The *Chainlit2*.*9*.*6* package provides a user interface to interact with the main agent. The agentic framework implements the following six successive steps (Figure 1):

A. Determine risk factors: The user provides a textual description of recent social interactions during which they may have been infected. The 100 most similar events to this description are retrieved using the Euclidean distance in the embedding space. From these events, we identify the six factors that most influence the LSTM’s prediction, both positively and negatively, using a Local Interpretable Model-agnostic Explanations (LIME) text explainer. An initial probability of infection *P*_0_ is inferred with the LSTM model based on the user’s description.
B. Generate question: Agent 1 asks a question reflecting one risk factor from the list and concerning the information provided by the user.
C. Assess question redundancy: Agent 2 independently assesses the redundancy of the question generated by Agent 1 compared to the current conversation. This aims to avoid asking similar questions to the user. If the question is deemed redundant, the process loops back to step B. Otherwise, the question is posed to the user.
D. Get user’s answer: The information provided by the user is stored for future context.
E. Refine risk: Agent 1 summarizes the whole conversation with the user, in the style of the contact tracing notes. We compute a new probability of infection *P*_*k*_, at iteration *k*, based on this summary, using the LSTM. If | *P*_*k*_ *− P*_*k−*1_ | *< ϵ* = 0.05, we assume that the estimation of the risk of infection has converged. If convergence is reached or all risk factors have been used to generate a question, the process follows with step F. Otherwise, it loops back to step B.
F. Generate risk report: Agent 1 provides a report summarizing the risk factors to the user, together with a prediction of their final risk of infection and whether convergence was reached along the iterations. The convergence information provides an uncertainty metric to the user.

In this framework, Agent 1 handles the conversation with the user. The agent is an instance of the pre-trained *Llama3*.*3 (70B)* model. To achieve the agentic behavior described above, we designed distinct prompts and supplied the agent with a specific context at steps B., E., and F. We could not predict the infection risk with this language model since this would require conversation data between the contact tracer and the user, which was not recorded. We therefore trained an independent supervised learning model for risk prediction. At step E., we provided a few-shot example to the agent in order to summarize the conversation according to the writing style of the contact tracing notes. This aims to infer predictions on textual data with a similar writing style as the ones used for training.

### 2.3 Optimizing the RAG framework

We aimed to optimize the conversational framework on five variables; the foundation model (*Mistral:7B, Gemma (12B), Llama3*.*3 (70B)*); the embedding model (*Gemma (7B), Nomic Embed (137M), Mxbai Embed Large (335M), Embedding Gemma (300M), Qwen3 Embedding (8B)*); the supervised learning model (Logistic regression, LSTM); the presence of the redundancy assessor; the retrieval heuristic (50 similar events, 100 similar events, fixed similitude threshold). We used three categories of metrics: LLM-as-a-judge metrics (Faithfulness, Relevance, Clarity, Naturalness); traditional machine learning performance metrics, like the ROC AUC, precision, recall, and F1-score; and various deterministic metrics to monitor the system’s behavior. Some metrics computed by the LLM-as-a-judge were provided by the Retrieval-Augmented Generation Assessment (RAGAs) framework [35], while others were computed based on a custom prompt. The metrics aimed to evaluate the text generated by Agent 1 (questions, summaries), the risk of infection predicted by the supervised learning techniques, the convergence of the agentic framework, the redundancy of the questions asked, and the retrieval step.

To evaluate the performance of our framework, we separated the set of events recorded by contact tracers into a training and a testing set. We solely used the former dataset to train the LSTM and generate the context of the RAG framework. For each foundation model, we simulated four series of 100 different conversations. In each conversation, the user was simulated with an independent pre-trained *Llama3*.*3 (70B)* instance. For each conversation, we provided the agent with a social description randomly sampled from the testing set as context. We designed the prompt so that this agent answers questions based on the content of the context, and replies “I do not know” when the answer cannot be inferred from it. Each series corresponds to a different training set and a different testing set. The initial event descriptions provided by the simulated user were sampled from the testing set. All the experiments were performed on a high-performance computing infrastructure maintained by the Flemish Supercomputer Center (Supplementary Table 1).

### 2.4 Evaluating the risk refinement of the RAG framework

We aimed to assess whether the conversational agent correctly stratifies users by their risk of infection. We simulated four synthetic user profiles with *Llama3*.*3*. These represent different contact patterns: a low-contact profile (few social interactions), a high-contact profile (frequent and diverse interactions), a routine profile (recurring contacts in stable settings such as auditoriums and households), and a disrupted-routine profile (atypical contacts associated with travel).

For each archetype, we described plausible social interactions happening in our student population during the pandemic. We separated the social interactions during the week and during the weekend to reflect students’ patterns. The simulated users were prompted to only answer questions asked by our conversational framework if they find the answers within our descriptions, and to reply “I do not know.” otherwise. We evaluated each profile by training four supervised learning models (i.e., four train/test splits). We simulated 100 conversations for each split and computed the average metrics over each set of 100 conversations.

As a first analysis, we recorded the probability of infection, as estimated by the LSTM at the end of the conversation between the conversational agent and the simulated user. We then computed the average probability over the 100 conversations for each archetype. We assessed whether those probabilities were discriminative between the different archetypes. For this purpose, we computed a pairwise discrimination ROC AUC metric. In this analysis, the ROC AUC does not reflect the predictive performance of the model, but rather quantifies whether the risk predictions differ between the simulations of the different archetypes. We computed this metric for each pair of archetypes. For each pair, we attributed a binary reference label: 0 for an archetype considered at low risk and 1 for an archetype considered at high risk. Based on those binary reference labels and the estimated probabilities of infection of each of the two archetypes, we then computed a ROC AUC for each pair. Finally, for each pair of archetypes, we tested whether their distributions of infection probability differed using a two-sided Mann–Whitney U test.

## 3 Results

### 3.1 Supervised learning

After validation of the hyperparameters, *RobBERT* achieved a mean ROC AUC of 0.73 (*Std* = 0.01), across 10 experiments. The logistic regression model trained on TF-IDF data and the LSTM model both performed similarly to *RobBERT*; they achieved a mean ROC AUC of 0.72 (*Std* = 0.01). However, scaling up to a much larger generative model did not improve predictive performance. A QLoRA-fine-tuned version of *Mistral NeMo* achieved a ROC AUC of 0.69 (*Std* = 0.02), slightly below the smaller models (Supplementary Table 2). *Word2Vec* preprocessing and gradient boosting classifiers did not show promising performance (Supplementary Figure 2).

For our conversational framework, neural network architectures are essential to capture language patterns (e.g., negative sentences) in summarized conversations with the user (Supplementary Figures 3, 4). Moreover, the difference in ROC AUC between the LSTM and *RobBERT* is small relative to the operational cost of the transformer. We therefore adopted the LSTM in the conversational framework, where inference latency at each turn is a practical constraint.

### 3.2 Agentic performance

We optimized the conversational framework with regard to different parameters. We observed different convergence rates and redundancy assessment rates between the open-source models built with 70 billion, 12 billion, and 7 billion parameters. We observed a higher convergence rate with language models with 70 billion and 12 billion parameters than with 7 billion parameters. On average, *Llama3*.*3* converges after a few more iterations than *Gemma3*. The proportion of questions judged redundant decreases with the complexity of the foundation model (Figure 2). Also, on average, foundation models of higher complexity generate questions and summaries of higher quality (Supplementary Figures 5, 6). Embedding contact tracing events with *Gemma (7B)* led to a high convergence rate and faster convergence than with *Mxbai Embed Large (335M), Nomic Embed (137M), Embedding Gemma (300M)*, and *Qwen3 Embedding (8B)*.

**Figure 1.**
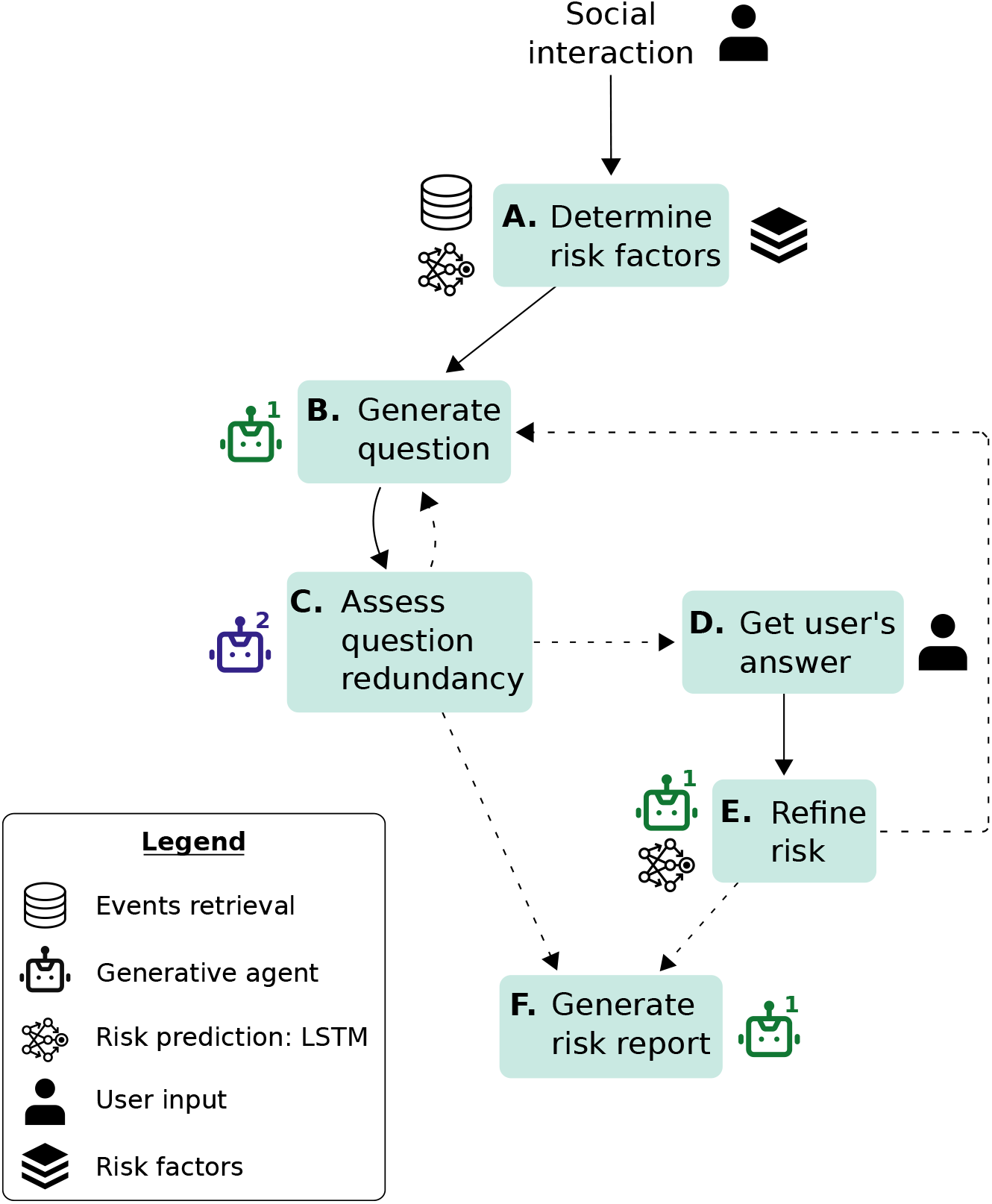
Architecture of the iterative RAG conversational agent. Within this graph, each node corresponds to an action implemented in the conversational framework. The directed edges determine the sequential behavior of the conversational system. The dashed edges represent conditional transitions to one node or another. Agent 1 iteratively collects information from the user and assesses their risk of infection with a pre-trained LSTM model. Agent 2 assesses the redundancy of the questions asked by Agent 1 to avoid asking similar questions to the user.

**Figure 2.**
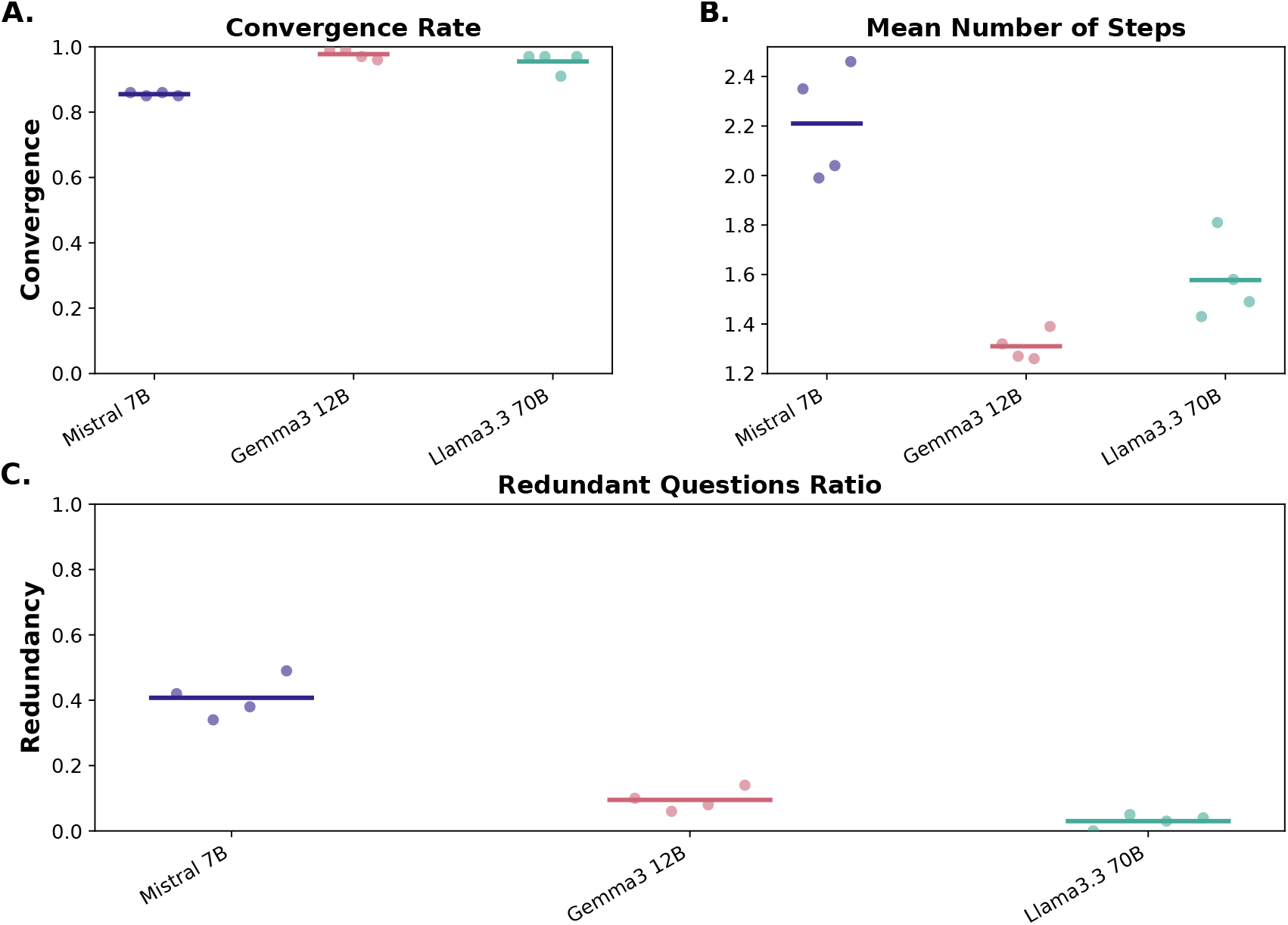
Impact of the foundation language model on the conversational framework. Each dot shows the average metric over 100 simulated conversations. Each bar shows the average between the four dots. (A, B) The conversational framework converged more often with *Llama3*.*3 (70B)* and *Gemma3 (12B)* than with *Mistral (7B)*. On average, *Llama3*.*3* required a few more steps than *Gemma3*. (C) While on average, 41% of the questions were deemed redundant with *Mistral*, only 9% of the questions were redundant with *Gemma3* and 5% of questions were deemed redundant with *Llama3*.*3*.

*Qwen3 Embedding* nonetheless remains a viable alternative, as it yielded higher RAG context relevance than the other models. (Supplementary Figures 7, 8). Retrieving the 100 most similar events to generate the list of risk factors seems to lead to a higher convergence rate (Supplementary Figures 9, 10). Moreover, the framework involving a redundancy assessment component requires fewer interaction steps with the user before convergence (Supplementary Figures 11, 12).

We also performed two additional experiments. First, we implemented a simple two-step RAG architecture that retrieves similar events and aggregates their risk labels to predict a binary risk of infection. Our analyses show that this architecture can slightly outperform the LSTM. However, it does not allow us to refine the risk of infection (Supplementary Figures 13, 14). Second, we compared the predictive performance of the LSTM on the summaries generated with and without a few-shot example. Including few-shot examples improved the prediction of infection risk (Supplementary Figures 15, 16).

The optimal conversational framework among the conducted experiments relies on *Llama3*.*3* (70B parameters), embeds the social descriptions with *Gemma* (7B parameters), retrieves the 100 most similar events to the initial description to generate the list of risk factors, and involves a secondary agent to assess the question redundancy (Supplementary Table 3).

We assessed the discrimination of the infection risks estimated among the different archetypes. The average end-of-conversation infection probability was highest for the high-contact archetype, followed by the disrupted-routine, routine, and low-contact archetypes (Figure 3.A). Then, we computed the discrimination AUC for each pair of archetypes. The probabilities estimated between the high-contact archetype and the low-contact archetype seem to be the most discriminative (*AUC* = 0.76). Four pairs of archetypes seem to present slightly discriminative probabilities of infection: low contact - routine (*AUC* = 0.62); low contact - disrupted routine (*AUC* = 0.64); routine - high contact (*AUC* = 0.65); disrupted routine - high contact (*AUC* = 0.61). According to the Mann–Whitney U tests, infection probabilities differed significantly between archetypes for those pairs (*p <* 0.001).The low p-values reflect the high number of simulated conversations for each archetype. However, the estimated infection probabilities did not differ significantly between the routine and disrupted-routine archetypes (*AUC* = 0.53; Mann–Whitney U, *p* = 0.14) (Figure 3.B). The conversational framework seems to gain more information during the conversations simulated with the high-contact archetype. Also, the infection risk seems to slightly correlate with our own perception of risk (Supplementary Figures 17, 18). We discussed an example of a conversation between the main agent and a simulated user in Appendix 14 (Supplementary Figures 19, 20).

**Figure 3.**
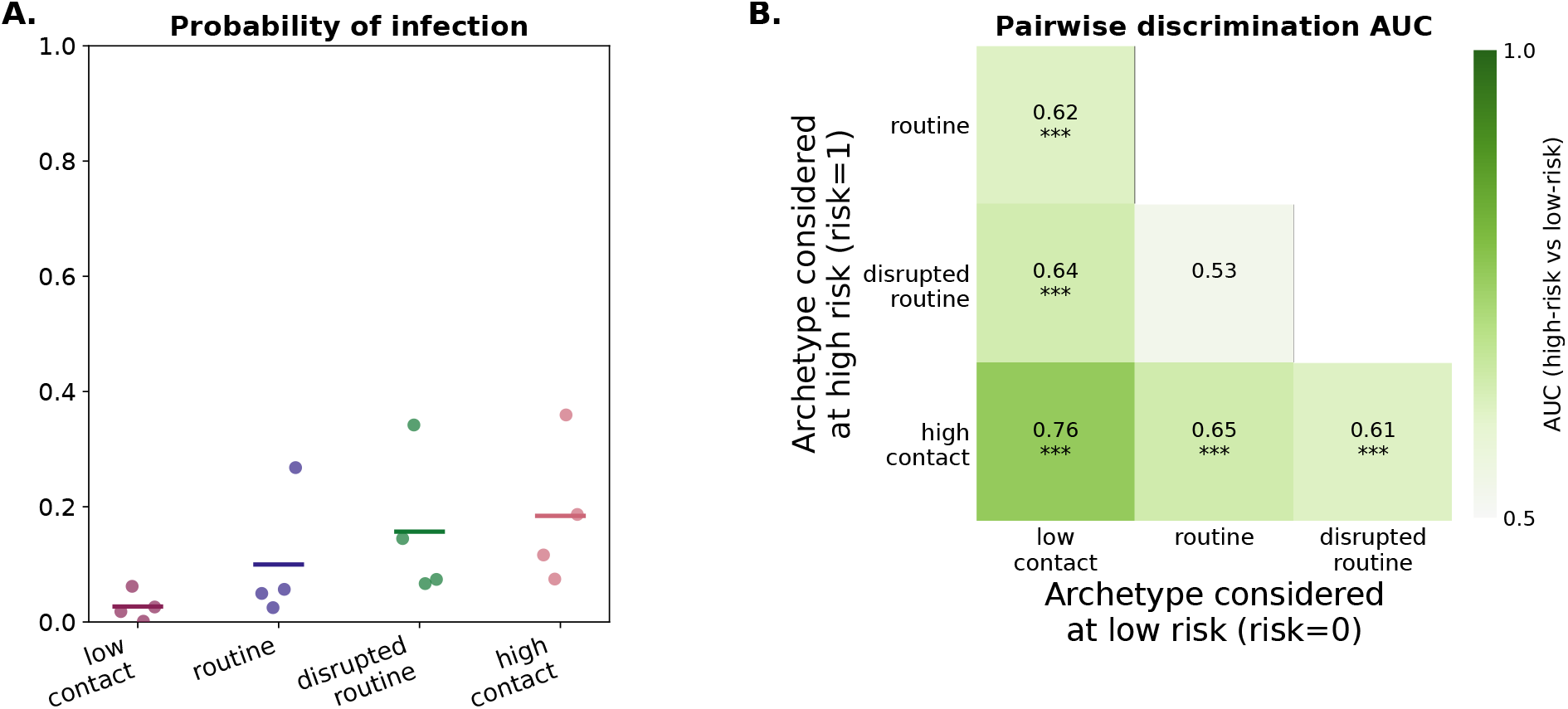
Discrimination of infection probabilities between the different archetypes. (A) Each dot shows the average probability of infection over 100 simulated conversations. Each bar shows the average between the four dots. The infection probability tends to decrease from the high-contact to the low-contact archetype (high-contact *>* disrupted-routine *>* routine *>* low-contact). (B) The ROC AUC values were computed based on all probabilities estimated for the different archetypes. The x-axis shows archetypes considered at low risk, while the y-axis presents archetypes considered at high risk for the AUC computation. ***: *p <* 0.001 (two-sided Mann–Whitney U test on the probability distributions of each archetype pair).

## 4 Discussion

Although LLMs have been used to predict the risk of infection [16, 17, 19, 20], to our knowledge, no study has aimed to predict infection risk from textual descriptions of social interactions. Contact tracing notes are precisely such descriptions, and predicting infection risk from them is our first goal. We implemented a RAG-enhanced framework to retrieve risk-labeled social interactions similar to an initial description of a social event provided by the user. As highlighted by Amugongo and colleagues, current RAG strategies deployed in healthcare lack iterative data refinement techniques. Therefore, as a second goal, we aimed to develop a conversational framework that asks relevant questions to the user to collect risk factors to iteratively refine the risk of infection [21]. Moreover, Belyaeva and colleagues emphasized the lack of explainability of conversational agent decisions [16]. Therefore, we controlled the questions iteratively asked of the user by generating a list of risk factors specific to the very first social interaction described by the user. In summary, in this study, we designed a hybrid conversational framework involving sequential steps together with agentic components to iteratively collect textual social context data from the user and refine their risk of infection. The supervised learning classifier shows good predictive performance. The optimal framework architecture presented promising generation metrics and a high convergence rate. Across simulated conversations, our system was able to differentiate the risk predictions between different social contexts.

To protect user privacy, the LLM inferences are performed on KU Leuven’s high-performance computing clusters and rely solely on open-source language models. Moreover, our framework allows us to iteratively refine the estimation of the infection risk by asking questions targeting possible risk factors for the user, relevant to their past conversation. This iterative process is informed by descriptions of social interactions during the COVID-19 pandemic and linked PCR diagnostics. Finally, our hybrid architecture offers both controllability over the agents’ behavior and the inherent autonomy of foundation language models. We can control the list of risk factors guiding the conversations based on public health experts’ opinions. Each component can also be adapted by modifying the prompts, for instance with few-shot examples, as demonstrated by our experiments on the summarization component. The relevance and naturalness of questions generated by the language model show a promising ability to converse with the user to investigate their recent social interactions. If implemented in a contact tracing procedure, our methods may help alleviate workforce requirements and may allow us to more rapidly highlight social context at risk of infection. These may help rapidly identify high-risk contacts of a positive case.

Nonetheless, our conversational framework presents some limitations. First, the context provided to the main agent solely relies on data collected during the COVID-19 pandemic in the student population in Leuven. The questions generated therefore reflect risk factors linked to a particular epidemiological context in a particular socioeconomic and sociodemographic group of the population [36]. The risk factors highlighted by the LSTM may have differed if the data had been sampled from another group of the population, or in the context of a different outbreak. Moreover, the textual content of the dataset influences both the predictive performance of the LSTM and the ability of the conversational framework to discriminate the infection probabilities between the users. Second, although open-weight foundation models ensure users’ privacy, proprietary language models could improve the overall generative performance of the framework, e.g., by asking more relevant questions to the user, given the past conversation. Third, we evaluated the conversational system based on simulated conversations. Indeed, since the conversational agent was not implemented during the pandemic period, our knowledge about the users’ social interactions is limited to data that were collected previously.

The conversational framework presented in this study may help identify contacts of positive cases at risk of infection. Targeting individuals at risk of infection based on the descriptions of their social interactions may optimize contact tracing. Further work could focus on fine-tuning LLMs based on actual contact tracing conversations led by public health professionals. With such methods, the generative models could ask relevant questions based on the provided training knowledge and therefore better refine the user’s infection risk, without requiring a separate supervised learning classifier. To this end, we suggest fine-tuning attention layers of different language models using LoRA algorithms [31, 32]. LLMs could also be useful for outbreak investigation. Such a tool could help identify outbreak-potential diseases and collect structured health data for epidemiological surveillance. For instance, we could collect symptom data and investigate possible exposure context to estimate incubation periods [37].

In conclusion, natural-language-based supervised learning techniques show good performance in predicting the individual risk of infection based on textual descriptions of social interactions. LLM-based agents could help refine the user-specific risk of infection, predicted based on their recent social interactions.

## Acknowledgments

The resources and services used in this work were provided by the VSC (Flemish Supercomputer Center), funded by the Research Foundation - Flanders (FWO) and the Flemish Government.

The authors acknowledge the assistance of Anthropic’s Claude Code technology (Opus 4.7) in providing coding and writing support. We also acknowledge the assistance of Consensus technology in describing the state of the art. All code and texts generated were reviewed and modified by the authors to ensure accuracy and functionality.

## Data availability

The analysis scripts are available at https://github.com/jothibaut/risk_prediction_chatbot.git. Individual-level health and social data cannot be shared to protect participant privacy and confidentiality.

## Authors’ Contributions

JT and JR directly accessed and verified the data reported in this article. All authors had full access to the study data, critically reviewed the manuscript, and approved the final version. They all accept responsibility for submitting for publication. JT contributed to the conceptualization of the study, the data curation, the analysis, the visualization, the programming, the methodology, and the writing of the original draft. MT, JDS, and JR contributed to the conceptualization of the study and the methodology. CG and LC contributed to the methodology. EA contributed to the methodology and the supervision.

## Ethical approval

The Ethics Committee Research UZ/KU Leuven approved the contact tracing study protocol (approval number S64919). Informed consent was waived, as data collection was limited to public health purposes.

## Abbreviations

CDC: Centers for Disease Control and Prevention
COVID-19: coronavirus disease 2019
ED: emergency department
GPT: Generative Pre-trained Transformer
LIME: Local Interpretable Model-agnostic Explanations
LLM: large language model
LoRA: Low-Rank Adaptation
LSTM: long short-term memory
NCD: non-communicable disease
PCR: polymerase chain reaction
QLoRA: Quantized Low-Rank Adaptation
RAG: retrieval-augmented generation
RAGAs: Retrieval-Augmented Generation Assessment
ROC AUC: area under the receiver operating characteristic curve
TF-IDF: term frequency–inverse document frequency

