## Appendices for "Using iterative agentic retrieval-augmented generation to refine the risk of infection from social contexts during the COVID-19 pandemic: Development and Evaluation Study"

#### Appendix 1 Data set description

The conversational framework described in the main analysis involves a database of descriptions of social interactions recorded within the student population in Leuven. Those descriptions were either reported by contact tracers or by the individuals themselves. In the main analysis, we solely focused on contact tracing data since risk prediction based on self-reported data achieved low performance (Supplementary Figure 2). Among the remaining social interactions, we excluded those without any positive cases. Those probably reflect missing information within our database, since contact tracing follows up a positive PCR. We considered that a social interaction was at risk (True label) if at least two participants tested positive within 14 days after the interaction. We considered the interaction at no risk (False label) if only one participant tested positive, i.e., if no pathogen transmission was recorded.

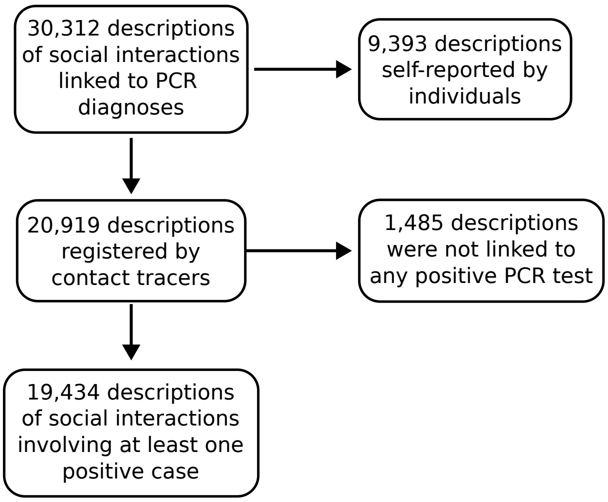


Supplementary Figure 1: **Data exclusion criteria.** 30,312 descriptions of social interactions involving participants with a known PCR diagnosis happened between October 5, 2020 and May 19, 2022. Among those, we retained 20,919 descriptions, recorded by contact tracers. We retained 19,434 descriptions of interactions involving individuals with at least one positive test.

#### Appendix 2 Computational architecture used to conduct the experiments

All experiments presented in this study were conducted on KU Leuven's high-performance computing infrastructure maintained by the Flemish Supercomputer Center (VSC). We distributed the experiments across different nodes, equipped with the following components (Supplementary Table 1) [1].

**B200:**

- 2x AMD EPYC 9655 CPUs (Turin), 96 cores each; frequency of 2.6 GHz.
- 1536 GiB RAM (default allocation of 8000 MiB per core).
- 8x NVIDIA B200 SXM6 (Blackwell) GPUs, 192 GiB GDDR, interconnected via NVLink.
- 960 GB NVMe local disk.

**H100:**

- 2x AMD EPYC 9334 CPUs (Genoa), 32 cores each (4 NUMA domains and 4 L3 caches per CPU); base and maximum CPU frequencies of 2.7 GHz and 3.9 GHz, respectively.
- 768 GiB RAM (default allocation of 11700 MiB per core).
- 4x NVIDIA H100 SXM5 GPUs, 80 GiB HBM3, interconnected via NVLink.

- 960 GB local SSD.

**A100:**

- 2x Intel Xeon Platinum 8360Y CPUs (Ice Lake), 36 cores each (2 NUMA domains and 1 L3 cache per CPU); Frequency of 2.4 GHz.
- 512 GiB RAM (default allocation of 7000 MiB per core).
- 4x NVIDIA A100 SXM4 GPUs, 80 GiB GDDR, interconnected via NVLink.
- 960 GB local SSD.

| Experiment | Node | Duration |
|---|---|---|
| Reference | H100 | 21h08 |
| Llama3 (70B, constrained) | B200 | 17h12 |
| Mistral (7B) | H100 | 22h57 |
| Gemma3 (12B) | B200 | 14h12 |
| Nomic Embed (137M) | H100 | 23h46 |
| Mxbai Embed Large (335M) | B200 | 20h55 |
| Embedding Gemma (300M) | H100 | 20h02 |
| Qwen3 Embedding (8B) | H100 | 21h52 |
| Retrieval - 50 | B200 | 18h07 |
| Retrieval - similarity | A100 | 26h56 |
| No redundancy grader | B200 | 16h42 |
| Logistic regression | B200 | 14h16 |
| No few-shots summary | B200 | 18h58 |
| 2-steps RAG | H100 | 00h35 |

Supplementary Table 1: **Duration and distribution of the experiments on the computing nodes.** The reference experiment refers to the framework presented in the main study. This framework uses *Llama3.3 (70B)* as a foundation model, *Gemma (7B)* as an embedding model, retrieves the top 100 events, uses a second agent to assess question redundancy, and uses LSTM as a prediction model (Supplementary Table 3). "Llama3.3 (70B, constrained)" refers to the use of this foundation model with a limit of *Ollama* context to 4,096 tokens. For this experiment, the number of generated tokens was limited to 200, 512, and 64 tokens for the simulated user, main agent, and redundancy grader, respectively. This experiment ensures a fair comparison with the *Gemma3 (12B)* and *Mistral (7B)* foundation models, which were also evaluated under those constraints, as described in Appendix 6.

### Appendix 3 Predicting the risk of infection with supervised learning techniques

As a first analysis before implementing the conversational framework, we compared the risk prediction performance between an LSTM architecture [2, 3], *XGBoost* [4, 5], and a simple logistic regression model [6]. For the last two models, we compared the predictive performance after pre-processing the textual data set with the TF-IDF algorithm and with *Word2Vec* [7, 8, 9]. We also compared the predictive performance of those models solely based on textual data describing social interactions, and by considering the week of the year during which it happened (integer $\in \{1, 53\}$) and the type of event ($\in$ {work environment, academic context, drinking establishment, dining establishments, own student residence, at friends' places, youth activity, family settings, travel context, private or public transport, other}). We also compared model performance when training on descriptions written by contact tracers, by the users themselves, or by both. Finally, we considered two different risk labels as ground truth to predict the risk of infection. The first label assumes that the social interaction described is at risk if at least one participant tested positive. The second label assumes that the social interaction described is at risk if at least two participants tested positive. The first risk label relies on the assumption that one single infected participant in a social event could transmit the pathogen to others. The second risk label relies on the assumption that two infected persons attending a social event may represent a transmission event, although the transmission hypothesis may not always be validated [10].

Supplementary Figure 2 shows the comparison of the ROC AUC obtained across all those experiments. We achieved higher performance with models trained on contact tracing data than with models trained on self-reported data. The week variable only slightly improves the predictive performance. The LSTM architecture and the logistic regression model trained on TF-IDF data present similar performance [2, 3, 7, 8]. They seem to perform better than *XGBoost*, and they performed better than predictive models trained on a data set vectorized with *Word2Vec* [4, 11, 9]. In the implementation of the conversational framework, we therefore retained the LSTM neural network, trained on unstructured textual data only. The memory components of this neural network account for natural language patterns over N-grams (e.g., negative sentences, adverbs, ...).

We also evaluated the predictive performance of transformer-based architectures, in comparison with the LSTM architecture. However, we did not assess their performance with data reported by participants themselves, nor while adding the type of event and the week of the year during which it happened. Instead, in those experiments, we solely focused on textual data sets, entirely collected by contact tracers, as selected from the experiments presented in Supplementary Figure 2. Transformer-enhanced neural networks do not seem to outperform the LSTM architecture (Appendix 5).

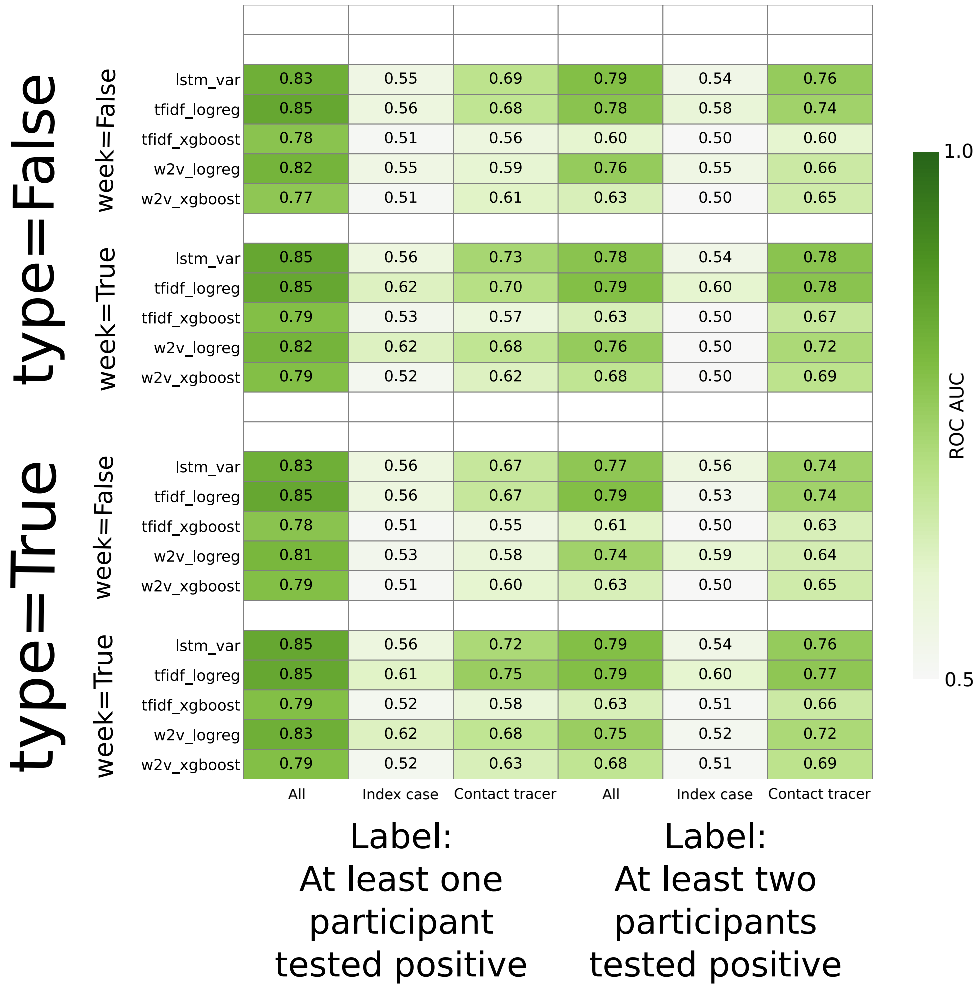


Supplementary Figure 2: **Risk prediction performance of different supervised learning models.** The predictive models trained on events collected by contact tracers showed higher performance than the ones reported by index cases. Moreover, the LSTM model often seems to outperform the other models, although the simple logistic regression model trained on TF-IDF data performed with a similar ROC AUC [7, 8]. Including the event's week or the type of event as a variable does not drastically improve the performance of the models. Finally, we achieved a higher ROC AUC with the ground-truth label reflecting the attendance of two infected persons to a social event.

### Appendix 4 Assessing the performance of the conversational framework across supervised learning models

To optimize the conversational framework, we evaluated its performance on different parameters. This section presents the impact of the supervised learning model used in the framework on its general performance. Appendix 6, Appendix 7, Appendix 8, and Appendix 9 present the impact on the framework performance of the foundation model, the embedding model, the retrieval heuristic, and the presence of the redundancy grader, respectively. We evaluated the general performance of the conversational framework with different metrics.

We grouped four metrics into the set of generation-quality metrics: faithfulness, relevance, clarity, and naturalness of the text generated by the LLM agent. Those four metrics were computed based on the assessment of an LLM-as-a-judge, instantiated from the *Llama3.3 (70B)* architecture. We used those metrics to assess both the questions and the conversation summaries generated by the main agent in the framework. Those metrics are defined as:

- Faithfulness:
  - Question: How factually consistent is a question with the previous summaries, and given a specific risk factor?
  - Summary: How factually consistent is a summary with the past conversation?
- Relevance:
  - Question: Is the question relevant to the specific risk factor and the previous conversation?
  - Summary: Does the summary accurately capture the key information from the conversation?
- Clarity: Is the generated text clear and easy to understand?
- Naturalness: Does the generated text sound natural in Dutch?

Additionally, we computed different metrics to evaluate the performance of the conversational framework at different steps:

- Prediction: We computed the ROC AUC based on the probability of infection computed by the supervised learning model, both at the beginning (initial event description) and at the end of the conversation, after refinement.
- Convergence: We computed two different metrics to evaluate the convergence.
  - Convergence rate: Proportion of conversations during which the convergence of the risk of infection was reached.
  - Mean number of steps: Among the conversations, the mean number of question-answer interactions between the bot and the user. The number of questions is limited to 6.
- Redundancy: We computed the proportion of questions asked by the main agent that were assessed as redundant by the redundancy grader.
- Retrieval: We computed two different metrics to evaluate the retrieval part.
  - RAG context relevance: Are the retrieved social events pertinent to the initial event description, which was randomly sampled from the testing set.
  - RAG label agreement: Among the retrieved social events, the proportion whose risk label matches that of the initial event description (randomly sampled from the test set).

The relevance, the clarity, and the naturalness were evaluated with our own custom prompts, while the faithfulness and the RAG context relevance were evaluated based on the RAGAs framework [12].

The risk prediction is estimated with a supervised learning model. According to the performance results presented in Appendix 3, considered outside our conversational framework, the LSTM model and the logistic regression (LR) model trained on TF-IDF data perform similarly and outperform XGBoost and TF-IDF trained on Word2Vec's output data [9, 4, 5, 7, 8]. As expected, the classifier model does not strongly influence the generation quality metrics (Supplementary Figure 3). The framework implemented with the TF-IDF LR model reaches convergence a bit faster than the one implemented with the LSTM model. However, the LR model reaches convergence less often. Also, the ROC AUC tends to decrease less with the LR model over the iterations. This can be explained by the inherent nature of the logistic regression algorithm; with this algorithm, prediction is solely influenced by the presence of words in the summary. If no word with a high predictive influence are generated from one step to another, the probability of infection does not vary a lot. Also, this model is limited since it does not account for negative sentences, nor other sequences of words. LSTM therefore performs better than LR for our conversational framework (Supplementary Figure 4). Fine-tuning transformer-based neural networks did not seem to improve the ROC AUC and required more time than LSTM. We therefore did not investigate their influence on the different generation and monitoring metrics (Appendix 5).

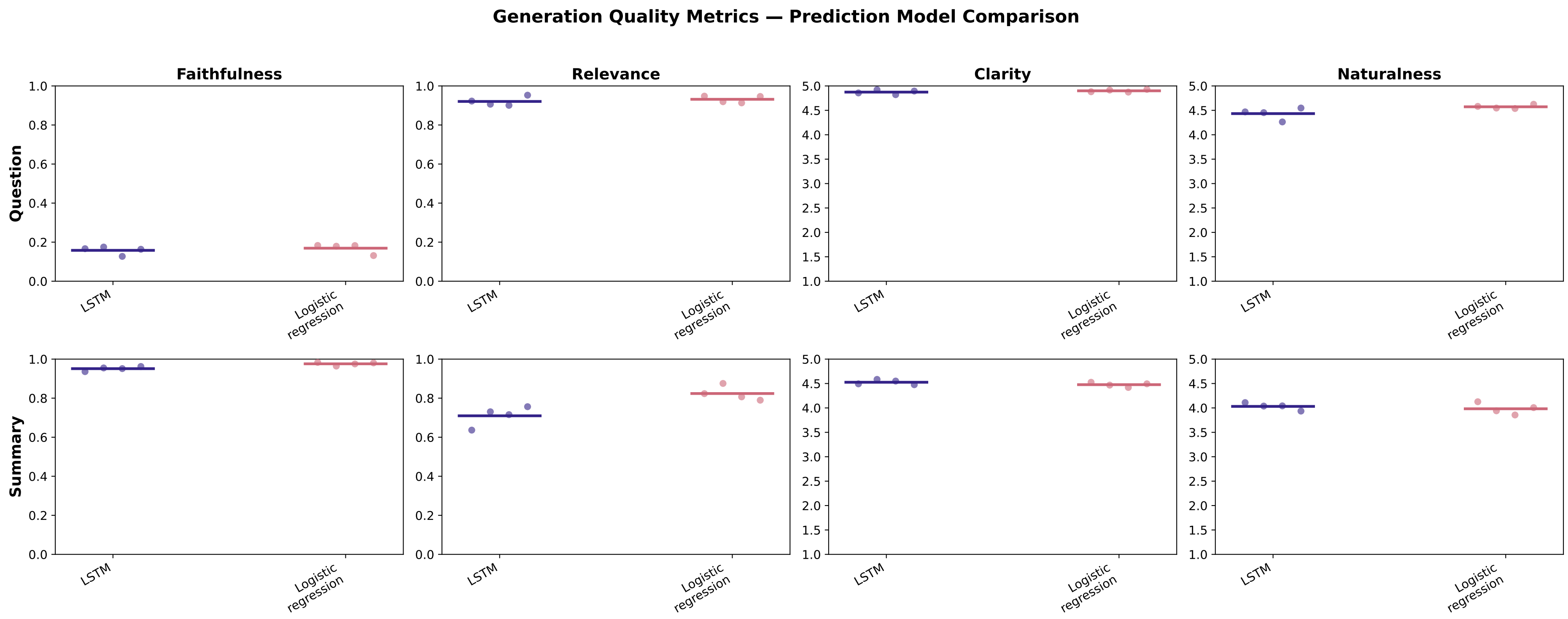


Supplementary Figure 3: **Comparison of the generative performance between an implementation with an LSTM model and an implementation with a logistic regression model.** The choice of classifier within our conversational framework does not strongly influence the generative performance of the model.

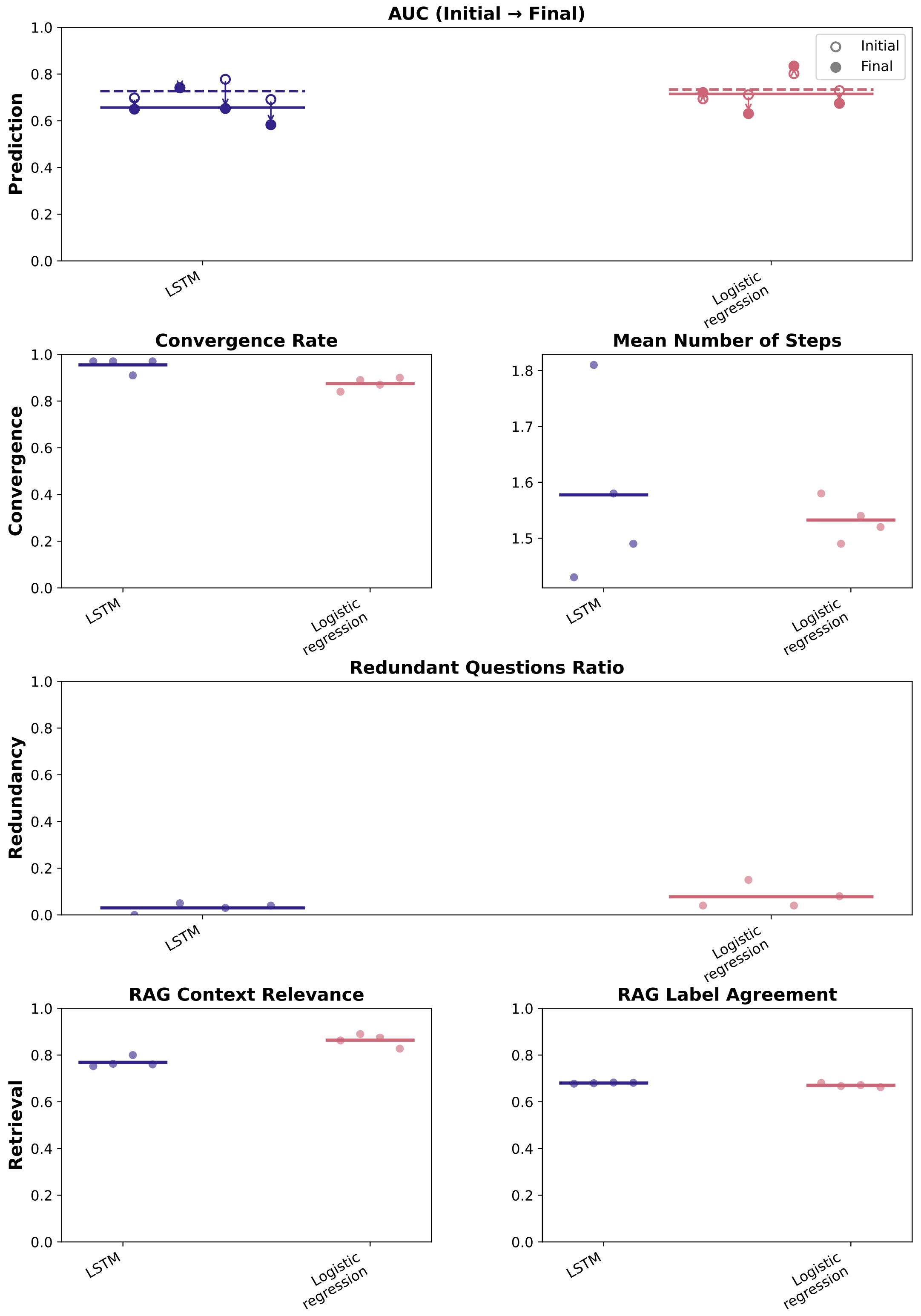


Supplementary Figure 4: **Comparison of the performance of the iterative agentic RAG framework at different steps between an implementation with an LSTM model and an implementation with a logistic regression model.** Although an implementation based on an LSTM model tends to decrease the AUC more over the iterations, it converges a bit more often and, on average, more questions are asked to the user. This can be explained by inherent algorithmic differences between those two models; while LR trained on TF-IDF [7, 8] data solely relies on the presence and frequency of words within a summary, LSTM considers sequences of words. Thus, it is more prone to highlight language nuances within a sentence and therefore refine the risk estimations.

### Appendix 5 Fine-tuning transformers to predict the risk of infection

Appendix 3 presented a comparison of the predictive performance between traditional natural language processing techniques and classifiers in predicting the risk of infection based on our dataset. We provided an estimation of the performance of the model based on different ground-truth labels, based on data gathered from different sources, and by additionally considering other predictive variables. In this section, we selected the two most promising simple models (logistic regression on TF-IDF, LSTM) and compared their performance with transformer-based architectures. We fine-tuned *RobBERT*, a Dutch RoBERTa-based language model [13], and *Mistral NeMo* (12B parameters) with QLoRA. We selected the latter model because it is multilingual, and fine-tuning can be performed on an 80GB GPU with low-rank adaptation [14, 15, 16].

To compare the performance of all the models, we first validated the hyperparameters of the LSTM architecture, *RobBERT*, and logistic regression. The QLoRA parameters were kept fixed at standard values [16]. Once the hyperparameters of all models were fixed, we tested each model on 10 different train/test splits of the dataset. We evaluated performance using the mean test ROC AUC across these splits, together with its standard deviation.

For the LSTM model, we validated the embedding dimension, the number of LSTM units, the learning rate, and the dropout rate. For the logistic regression, we validated the regularization strength, the penalty type ($\ell_1$, $\ell_2$, or none), and the class weighting. All parameters of the *RobBERT* sequence classification model - the pre-trained encoder and the randomly initialized classification head - were fine-tuned on our contact tracing dataset; we validated the learning rate, the batch size, and the weight decay.

In the *Mistral NeMo* architecture, we used QLoRA [16] to reduce memory requirements: the weights of the base model were quantized to 4 bits using the *NormalFloat4* data type with double quantization, and kept frozen, while computations were carried out in `bfloat16`. On top of this quantized backbone, we trained low-rank adaptation matrices (rank $r = 16$, $\alpha = 32$) on the four attention projections (query, key, value, output), together with a newly initialized classification head.

Based on our experiments, we observed that *RobBERT* performs similarly to the simpler LSTM and logistic regression models. However, those three models seem to slightly outperform *Mistral NeMo*. The encoder-only model retained an advantage on this small, Dutch-language classification task (Supplementary Table 2).

| **Model** | **Mean** | **Std** |
|---|---|---|
| *RobBERT* | 0.73 | 0.01 |
| Logistic regression | 0.72 | 0.01 |
| LSTM | 0.72 | 0.01 |
| *Mistral NeMo* (QLoRA) | 0.69 | 0.02 |

Supplementary Table 2: **Comparison of the predictive performance between four supervised learning models, after validation.** The four models achieved similar performance, with a mean ROC AUC between 0.69 and 0.73. LSTM, *RobBERT*, and the logistic regression slightly outperformed *Mistral NeMo* fine-tuned with QLoRA.

### Appendix 6 Assessing the performance of the conversational framework across foundation models

We compared the performance of our conversational framework based on three different foundation models: *Mistral* (7B parameters) [17], *Gemma3* (12B parameters) [18], and *Llama3.3* (70B parameters) [19]. For most experiments performed in this study (described in Appendix 7, Appendix 8, Appendix 9, Appendix 3, Appendix 12), we set the *Ollama*'s context window limit to 8,192 tokens. This allows the language model loaded by *Ollama* to attend to the full generated conversation, in particular when summarizing it.

However, for the configuration using the *Gemma3 (12B)* model for the main agent, this limit was occasionally exceeded. Indeed, the lower-quality questions generated by *Gemma3 (12B)* sometimes caused the simulated user (powered by *Llama3.3 (70B)*) to produce degenerate, repetitive answers spanning thousands of tokens, which in turn inflated downstream prompts beyond the 8,192-token budget. Also, we observed deadlocks for the configuration using *Mistral (7B)* with the 8,192-token context. Therefore, to prevent answer degeneration and deadlocks for the experiments with any of the three foundation models, we capped the agent's text generation at 512 tokens, the redundancy grader's generation at 64 tokens, and the simulated user's response length at 200 tokens. We assumed that relevant questions, redundancy assessment, and answers are necessarily shorter. Also, with those constraints fixed, we set the limit of *Ollama*'s context to 4096 tokens to accelerate the experiments. This limit was never exceeded over the experiments.

The performance of the conversational framework varies with the foundation model used to implement the agents. The questions and summaries often seem to present higher relevance, clarity, and naturalness when generated with foundation models designed with more parameters. However, *Llama3.3* is judged a bit less faithful than *Gemma3*. As expected, the faithfulness of the questions remains low for each foundation model, since the questions aim to gather new information, whose answers are unknown to the past conversation (Supplementary Figure 5).

The ROC AUC decreases from the risk predicted based on the initial social context to the risk estimated based on the refined summary generated by the agent. The descriptions used to train the LSTM and the initial event descriptions were all written by contact tracers, while the final summary is generated by an LLM. This may justify the low AUC at the end of the conversation. The agentic frameworks built with *Gemma3* and *Llama3.3* converge more often. On average, *Llama3.3* asked a few more questions before convergence. Questions generated by complex models seem to be graded as less redundant to the conversation. The event retrieval does not rely on the foundation models of the agents, but only on the embedding architecture and therefore shows low variability in this experiment (Supplementary Figure 6).

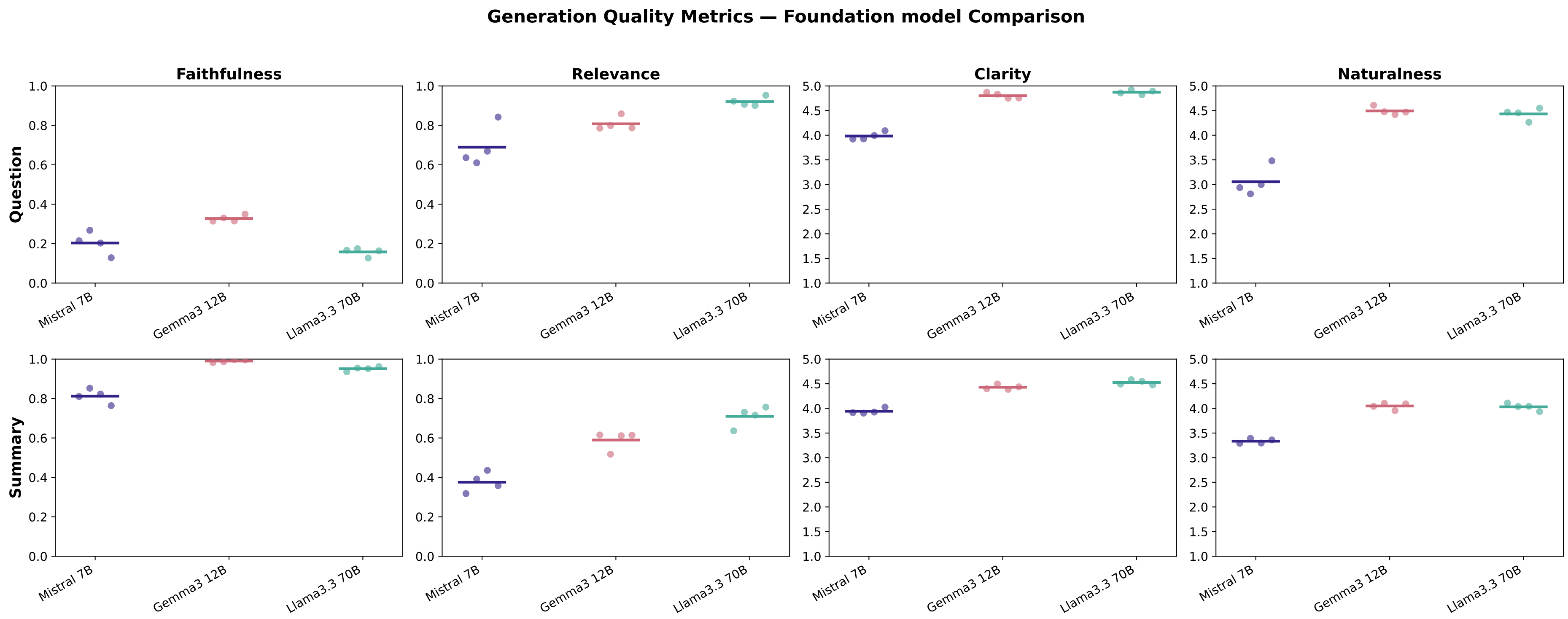


Supplementary Figure 5: **Comparison of the generative performance between implementations with three foundation models.** Each dot represents the average metric computed over 100 simulated conversations. The horizontal bars show the average metric between the four series of conversations (4x 100 conversations). The three foundation models are built respectively on 7 billion, 12 billion, and 70 billion parameters. The more complex models often show higher generative performance.

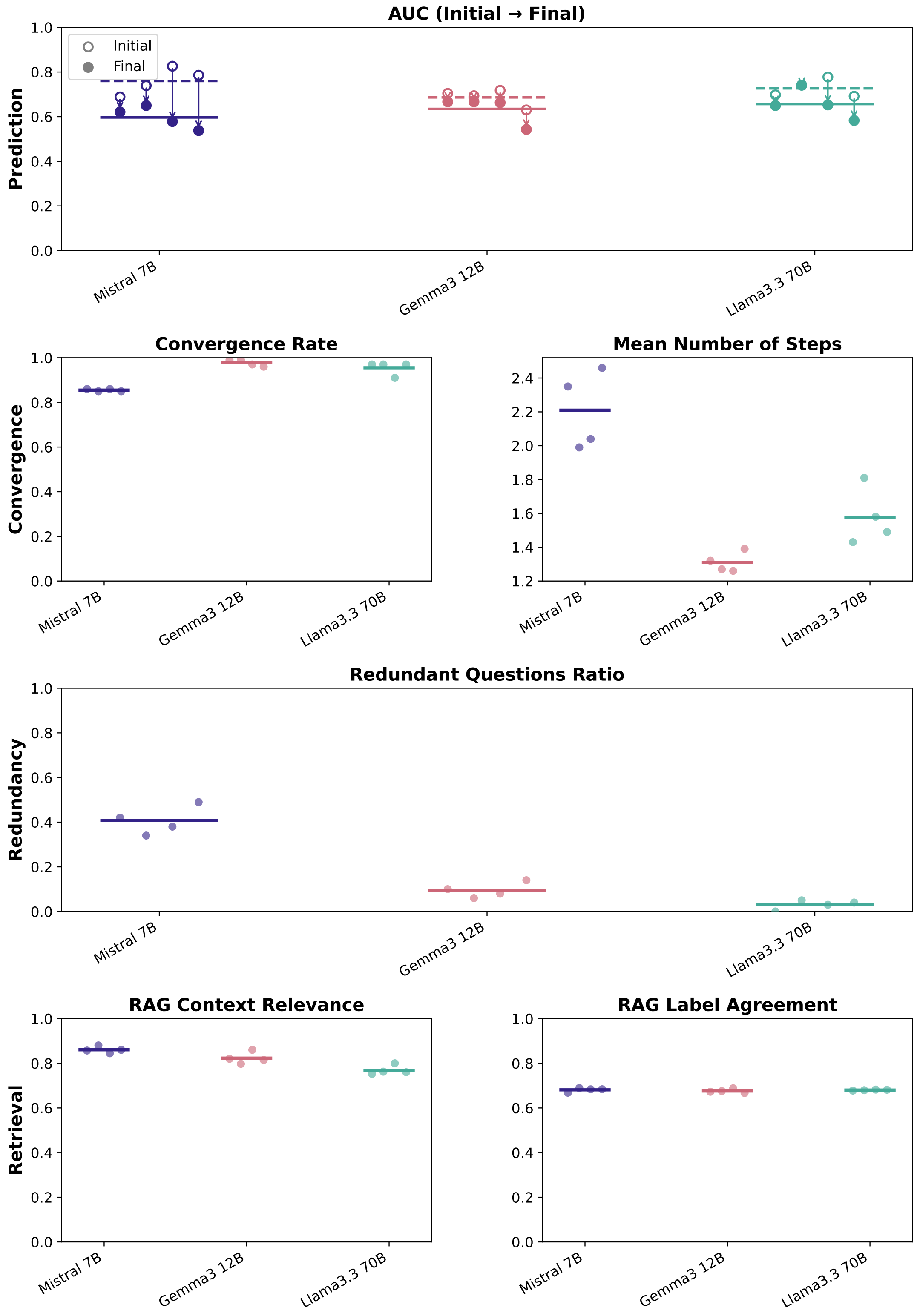


Supplementary Figure 6: **Comparison of the performance of the iterative agentic RAG framework at different steps between three foundation models.** While the ROC AUC of the LSTM decreases along the conversation, the system allows it to collect more information from the user. Increasing the complexity of the foundation model allows it to converge more often, with fewer redundant questions.

### Appendix 7 Assessing the performance of the conversational framework across embedding models

We studied the influence of five different embedding models on the performance of the conversational framework. We selected a sample of embedding models for comparison according to their number of parameters, their training objective, and their ranking on the Massive Text Embedding Benchmark (MTEB) (accessed June 30, 2026) [20, 21]. Moreover, to simplify the implementation, we restricted our selection to the embedding models supported by the *langchain_ollama1.0.1 Python* package. Specifically, we compared data embedding using *Gemma (7B)* with *Nomic Embed (137M)*, *Mxbai Embed Large (335M)*, *Embedding Gemma (300M)*, and *Qwen3 Embedding (8B)* [22, 23, 24, 25, 26]. *Gemma (7B)* is a generative model repurposed for embeddings, whereas the other four were trained specifically as embedding models. We included *Qwen3 Embedding (8B)* because it ranked fourth in the multilingual category on MTEB at the time of access, while its size remained small enough to be loaded together with *Llama3.3 (70B)* on a single GPU.

The choice of embedding model does not appear to strongly influence the generation-quality metrics. Since summary generation does not involve the embedding model, the apparent upward trend in summary relevance across models cannot be attributed to it and most likely reflects random variation (Supplementary Figure 7). Different embedding models may be preferred depending on the selection criteria. In the main study, we retained *Gemma (7B)* because it achieved a high convergence rate in fewer steps than the other models, averaged over our experiments, and because it produced few redundant questions. *Qwen3 Embedding (8B)* reached the highest RAG context relevance and could therefore also be a good fit for this application. Finally, a lighter model such as *Nomic Embed (137M)* achieved comparable performance while requiring considerably less memory, making it suitable for resource-constrained settings (Supplementary Figure 8).

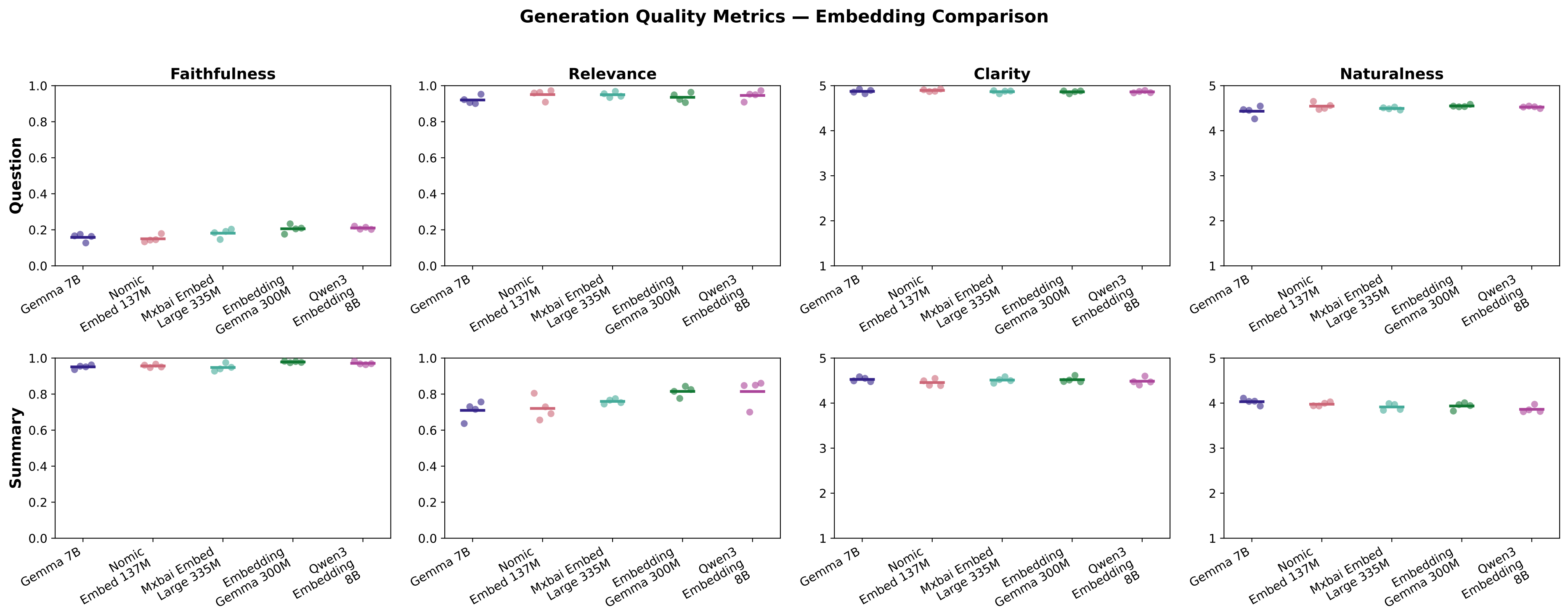


Supplementary Figure 7: **Comparison of the generative performance between five different embedding models.** The embedding model does not seem to strongly influence the generation quality metrics.

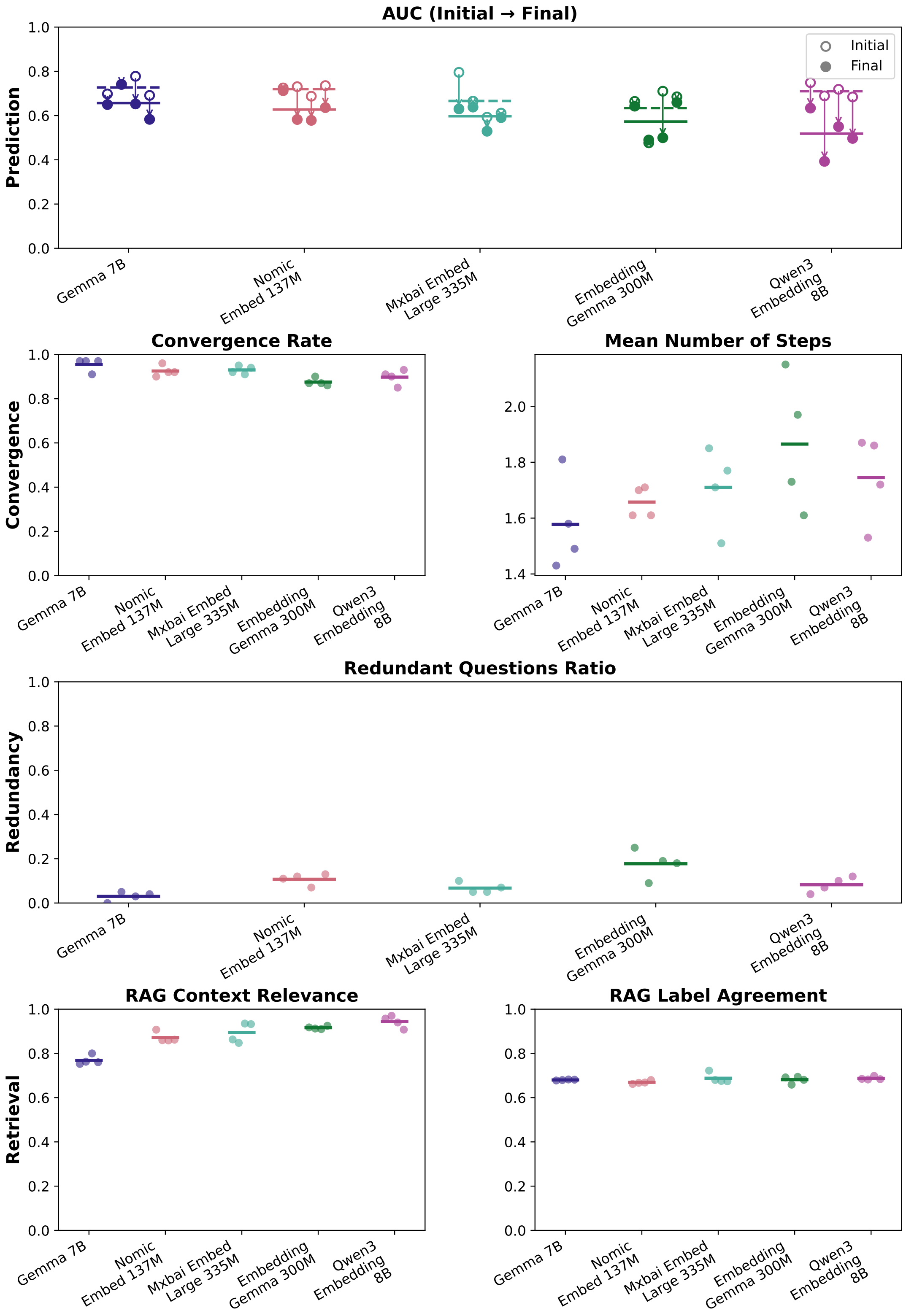


Supplementary Figure 8: **Comparison of the performance of the iterative agentic RAG framework at different steps between three embedding models.** With *Gemma (7B)*, the framework reached a high convergence rate in fewer iterations than with the other models. *Embedding Gemma (300M)* appears to produce slightly more redundant questions. The models trained specifically as embedding models yielded higher RAG context relevance than *Gemma (7B)*. With *Gemma (7B)*, the generated questions were slightly less redundant.

### Appendix 8 Assessing the performance of the conversational framework across retrieval heuristics

As a first interaction with the agent, the user describes the recent social interactions that they consider at risk. We then retrieve known event descriptions in our database that are similar to this initial description. A description is similar to another if the Euclidean distance between those descriptions is short in the embedding space. Among the events retrieved, we perform a LIME analysis to study the words that influence the LSTM model the most. Those words compose a stack of risk factors which serve the agent to generate questions.

In this section, we studied the impact of the retrieval heuristic on the performance of the conversational framework. We conducted three experiments: we first retrieved the 100 most similar events to the initial description, for the lime analysis; we then retrieved the 50 most similar events; we finally retrieved events based on a fixed similitude threshold. This similitude threshold is computed as the $20^{th}$ percentile among all pairwise distances within our embedding set. If more than 100 events were retrieved based on this similitude threshold, we only retained the 100 descriptions that are the most similar to the initial description for further analysis. This ensures that the prompt to compute the context relevance remains within the 8196-token limit.

As expected, the generation quality metrics are not influenced by the retrieval heuristic. Indeed, the retrieval heuristic influences the stack of risk factors, which should not impact the LLM generations (9). However, retrieving the 100 most similar events leads to the highest convergence rate and leads to faster convergence than with the 50 events retrieval. However, the top 100 events retrieved seem to be, on average, a bit less relevant to the initial event description than the top 50 events retrieved, or the events retrieved based on a fixed Euclidean distance threshold. Nonetheless, from those observations, the heuristic which retrieves the 100 most similar events seems to outperform the two other heuristics.

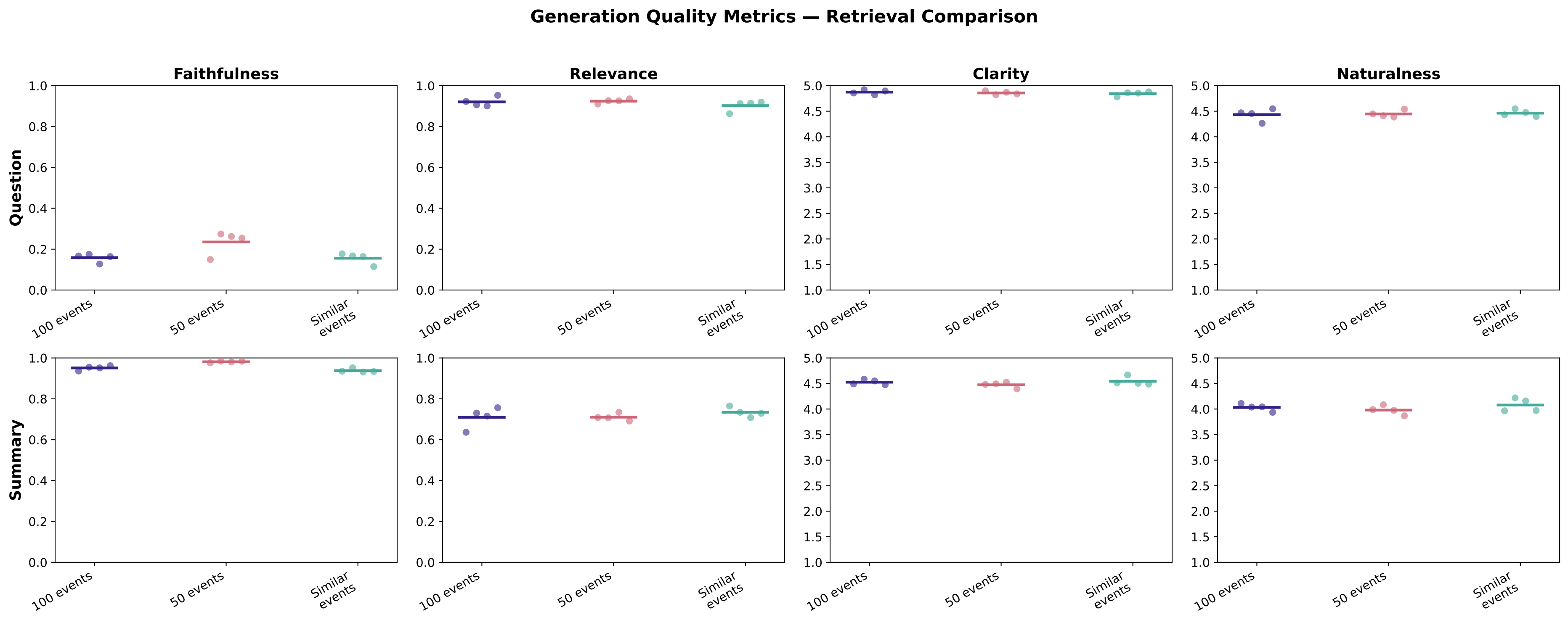


Supplementary Figure 9: **Comparison of the generative performance between three different retrieval heuristics.** We studied the impact of retrieving the 50 most similar events to the initial description, the 100 most similar events, or the events more similar than a fixed similitude threshold. The choice of the heuristic does not impact the generation quality metrics.

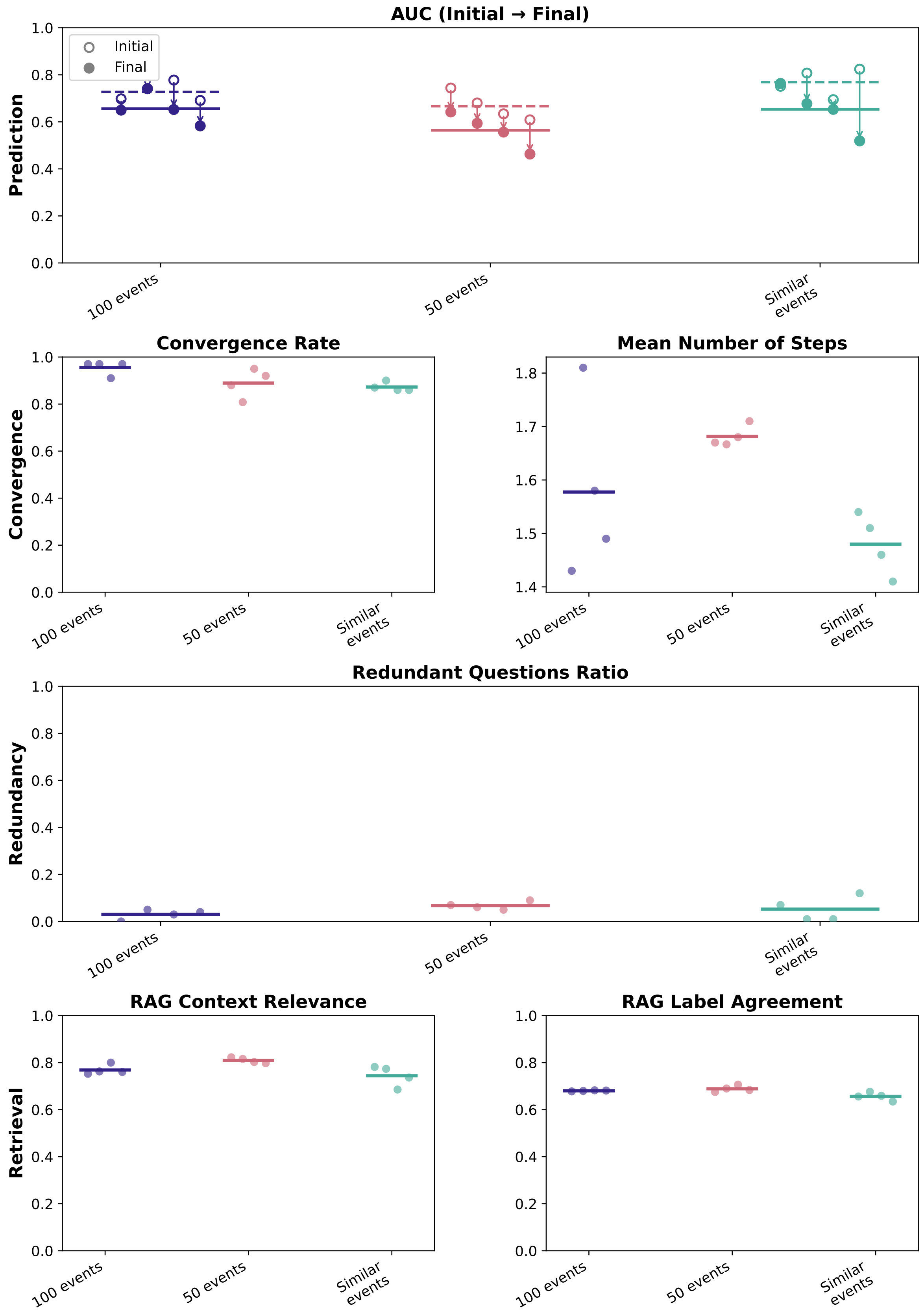


Supplementary Figure 10: **Comparison of the performance of the iterative agentic RAG framework at different steps between three different retrieval heuristics.** Retrieving the 100 most similar events to the initial user's description leads to a higher RAG rate of convergence than with the two other heuristics.

### Appendix 9 Assessing the performance of the conversational framework with and without the redundancy grader

The framework architecture involves two different LLM agents; one directly interacts with the user, and the other serves as a grader to assess the redundancy of a questio, compared to the previous conversation (Figure 1). We evaluated the impact of the redundancy grader on the performance of the conversational framework. The presence of the redundancy component does not seem to drastically influence the generative performance of the framework (Supplementary Figure 11). However, the redundancy assessor led to convergence in fewer iterations. The slightly lower RAG context relevance observed with the question redundancy component is likely due to random variation, since retrieval occurs before any redundancy check (Supplementary Figure 12). Those experimental observations support the reason for adding this component within the architecture of our framework. It indeed correctly fulfills its main goal in ensuring that the main agent asks unique, and therefore fewer, questions to the user.

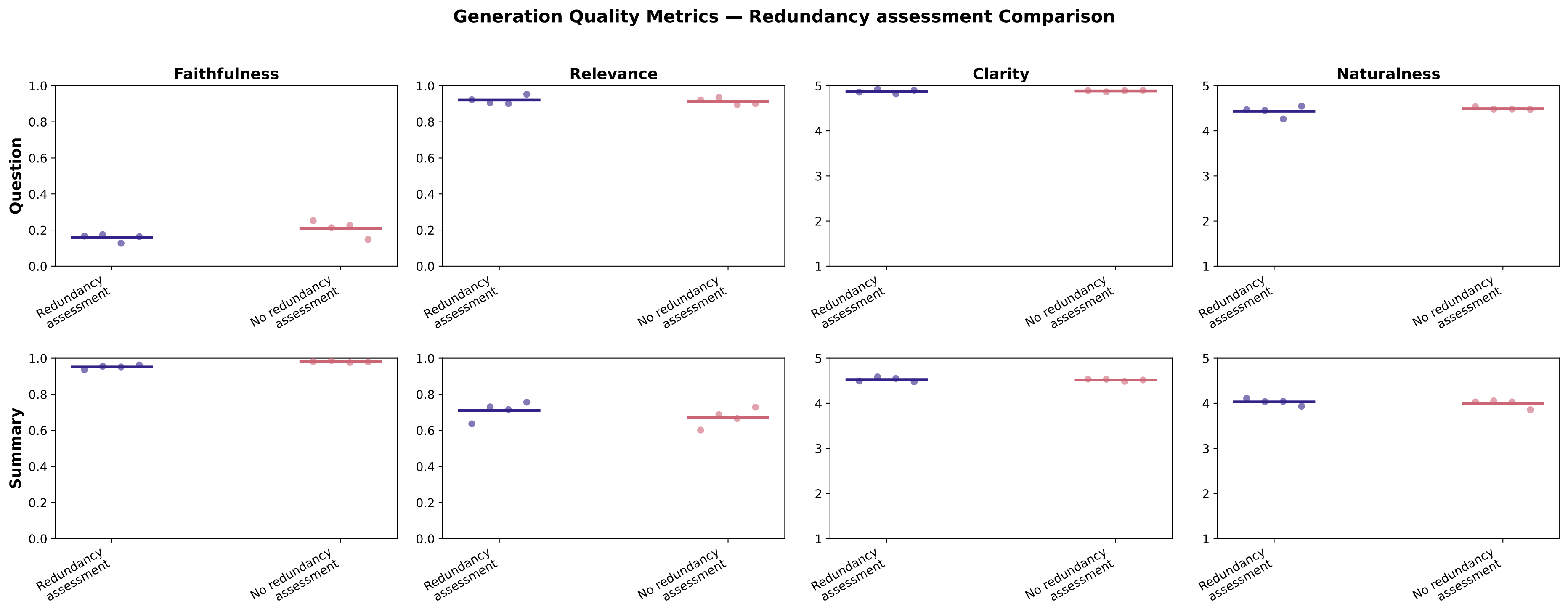


Supplementary Figure 11: **Comparison of the generative performance with and without the redundancy assessment component.** The presence of the redundancy assessment component does not seem to strongly influence the performance metrics.

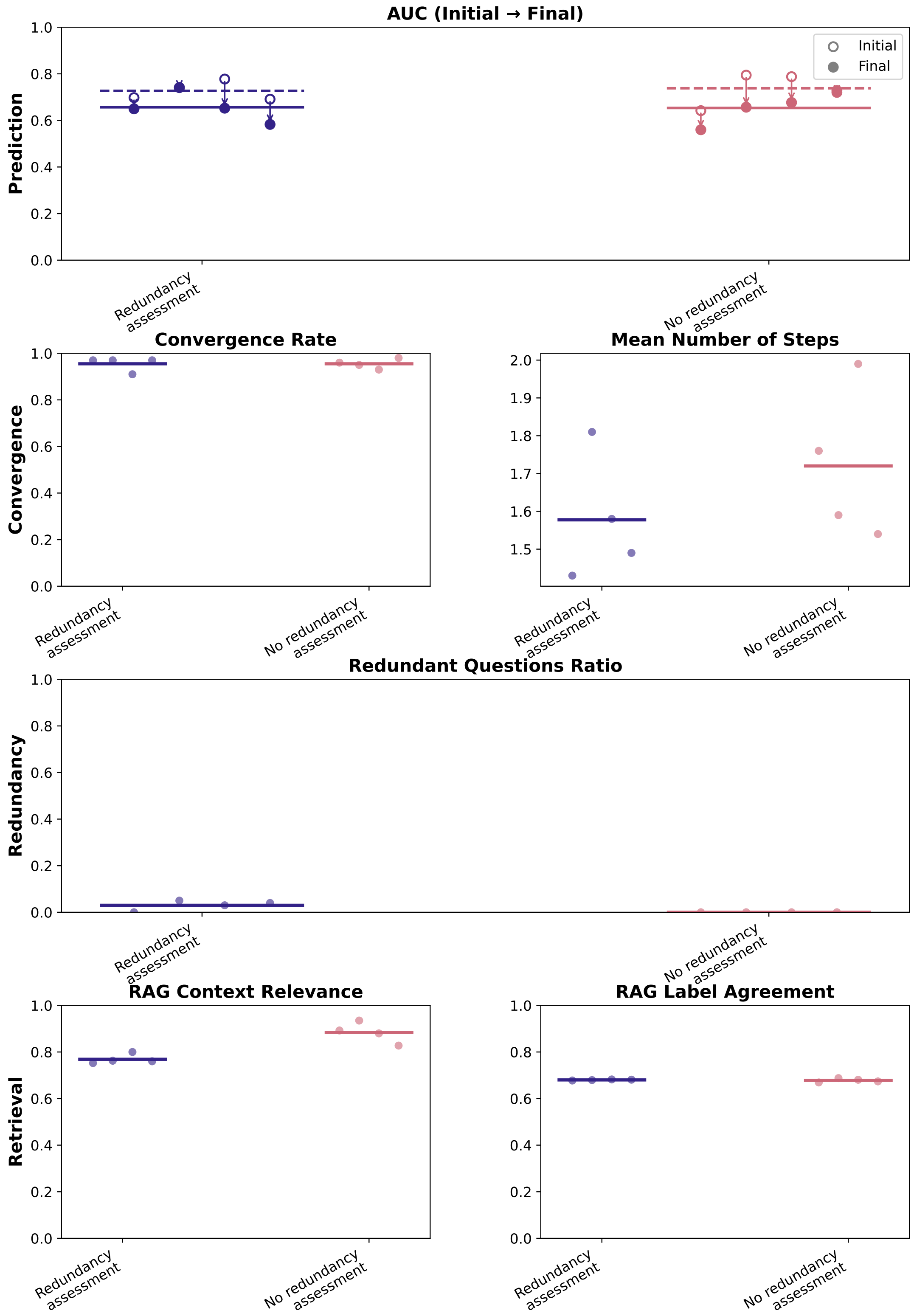


Supplementary Figure 12: **Comparison of the performance of the iterative agentic RAG framework at different steps with and without the redundancy assessment component.** The redundancy assessor met its objective since its presence led to convergence in fewer iterations.

### Appendix 10 Optimal framework

Based on the experiments described in Appendix 4, Appendix 6, Appendix 7, Appendix 8, Appendix 9, the framework that performed best among the ones explored relied on the Llama3.3 (70B parameters) foundation model [19]. Indeed, the high complexity of this LLM allowed it to generate high-quality questions and summaries. Embedding the data with Gemma (7B parameters) led to faster convergence. Retrieving the 100 most similar events to generate the stack of risk factors led to the highest convergence rate. Then, involving a redundancy assessment component allowed the framework to converge in fewer iterations. Finally, the LSTM model performed better than a logistic regression model; it led to a higher convergence rate, and it accounts for word sequences and local context. The suggested optimal conversational framework is highlighted in Supplementary Table 3, which compares all our experiments.

| Variable | Parameter 1 | Parameter 2 | Parameter 3 | Parameter 4 | Parameter 5 |
|---|---|---|---|---|---|
| Foundation model | Mistral 7B | Gemma3 12B | Llama3.3 70B | | |
| Embedding model | Gemma 7B | Nomic Embed 137M | Mxbai Embed Large 335M | Embedding Gemma 300M | Qwen3 Embedding 8B |
| Retrieval method | 100 events | 50 events | Similarity threshold | | |
| Redundancy grader | Present | Absent | | | |
| Prediction model | LSTM | Logistic regression | | | |

Supplementary Table 3: **Optimal framework for all conducted experiments.** Our experiments suggest that the best framework relies on the Llama3.3 (70B) foundation model and the Gemma (7B) embedding model [19]. It retrieves the 100 most similar events to the initial descriptions and relies on a second agent to assess the redundancy of the questions generated by the main agent. Finally, it predicts the risk of infection with an LSTM model. The parameters of the optimal architecture are highlighted in green.

### Appendix 11 Assessing a simple 2-step RAG

We compared the predictive performance of the LSTM architecture with a sequential 2-step RAG pipeline (Supplementary Figure 13):

A. We retrieve the 5 most similar events to the description of the social interaction provided by the user. Those are the 5 descriptions at the shortest Euclidean distance to the initial description in the embedding space. Embedding is performed with *Gemma (7B)*. Each retrieved event is linked to a risk label. A social event is considered at risk if at least two attendees had a positive PCR test.

B. We provide the initial social textual description to a *Llama3.3 (70B)* agent, together with the similar events and their risk labels [19]. Based on this information, the agent is prompted to assess a binary risk of infection for the user.

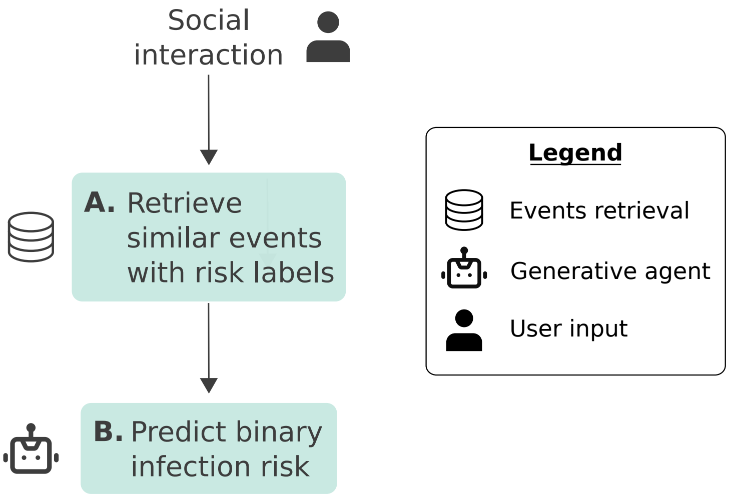


Supplementary Figure 13: **Architecture of the sequential 2-step RAG agent.** (A) The agent retrieves the 5 most similar events within the contact tracing data set, together with their corresponding risk labels, i.e., true if at least two attendants tested positive and false otherwise. (B) Based on the 5 events retrieved and their risk labels, Llama3.3 (70B) is prompted to assess whether the social description provided by the user presents a risk of infection, or not (binary prediction) [19].

Following the general methodology of the previous experiments, we evaluated four times 100 risk assessments for each model. We randomly distributed our data into a training set and a testing set. The initial social interactions were randomly sampled from the testing set, while the similar events retrieved belonged to the training set. Since the RAG prediction is binary, we compared its performance with the LSTM architecture using the precision (i.e., positive predicted value), recall (i.e., sensitivity), and $F_1$-score:

$$F_1 = 2 * \frac{Precision * Recall}{Precision + Recall}$$

The decision threshold for the LSTM was set at 0.5. The 2-step RAG slightly outperformed the LSTM. Indeed, it predicted the risk of infection with a higher $F_1$-score, reflecting a higher recall, although it presented a slightly lower precision on average. However, such an architecture does not allow us to iteratively refine the risk of infection by asking more questions about the user's social interactions. Its good performance solely reflects the risk predicted on one initial social description. Therefore, we still recommend using the optimal architecture presented in Appendix 10.

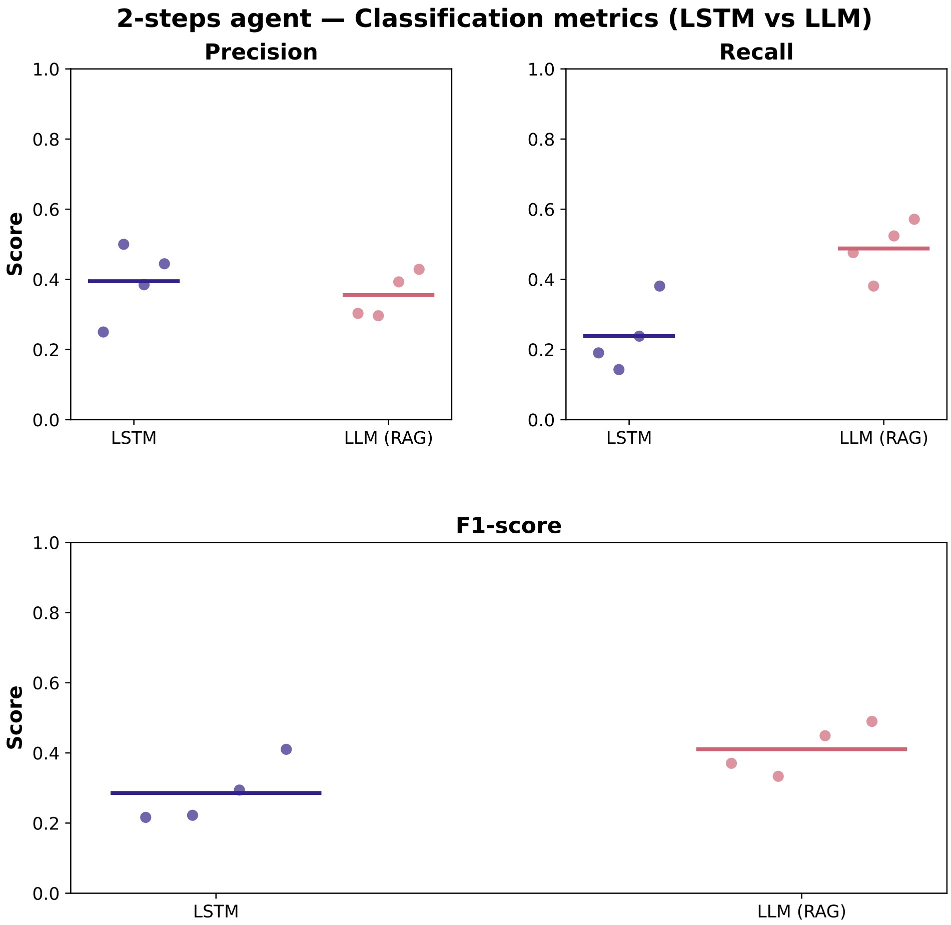


Supplementary Figure 14: **Comparison of the predictive performance between an LSTM model and a 2-step RAG implemented with Llama3.3 (70B) [19].** On average, the two models showed similar performance. The RAG model achieved a better $F_1$-score, reflecting a higher recall. However, the precision of the RAG seems to be slightly lower than the LSTM.

### Appendix 12 Influence of adding a few-shots examples in the summary prompt

We studied the performance of the conversational framework with two different prompts for the summarization component: a simple summarization prompt without any example, and a second prompt with a 5-shot example. The latter version of the prompt aims to generate a summary in a writing style similar to the notes taken by contact tracers, given a conversation between the agent and the user. Therefore, the LSTM classifier infers the risk of infection based on a textual description similar to the ones used for training. With the simple prompt, we frequently observed a drop in infection probabilities in the iterative process, leading to a rapid convergence towards a close-to-zero risk of infection. The improved prompt seems to solve this issue; the risk predictions seem to vary with the different risk factors across iterations. Therefore, in the main analysis, we used the summarization prompt with a few-shot example.

We compared the generative performance and monitored the behavior of the framework with both implementations of the summarization prompt. The summaries generated with the few-shot prompt are as faithful as those generated with the simple prompt. However, they lose relevance, clarity, and naturalness. Those last three metrics indeed reflect the writing style of contact tracing notes, which are usually very short and synthetic (Supplementary Figure 15).

Interestingly, the AUC tends to decrease much less over the iterations when following the writing style of contact tracers. This can probably be explained by the fewer steps taken during the iterative process with the enhanced prompt. Indeed, with this latter prompt, if the user answers “Ik weet het niet.” (i.e., ”I don’t know.”), the agent likely generates a summary similar to that of the previous step, which leads to estimating a similar probability of infection and therefore leads to convergence, i.e., the end of the process. In a real deployment, this behavior would likely be observed less often since the user would probably know the answer to the agent's questions. The few-shot example prompt did not influence the convergence rate, the questions' redundancy, nor the retrieval metrics (Supplementary Figure 16).

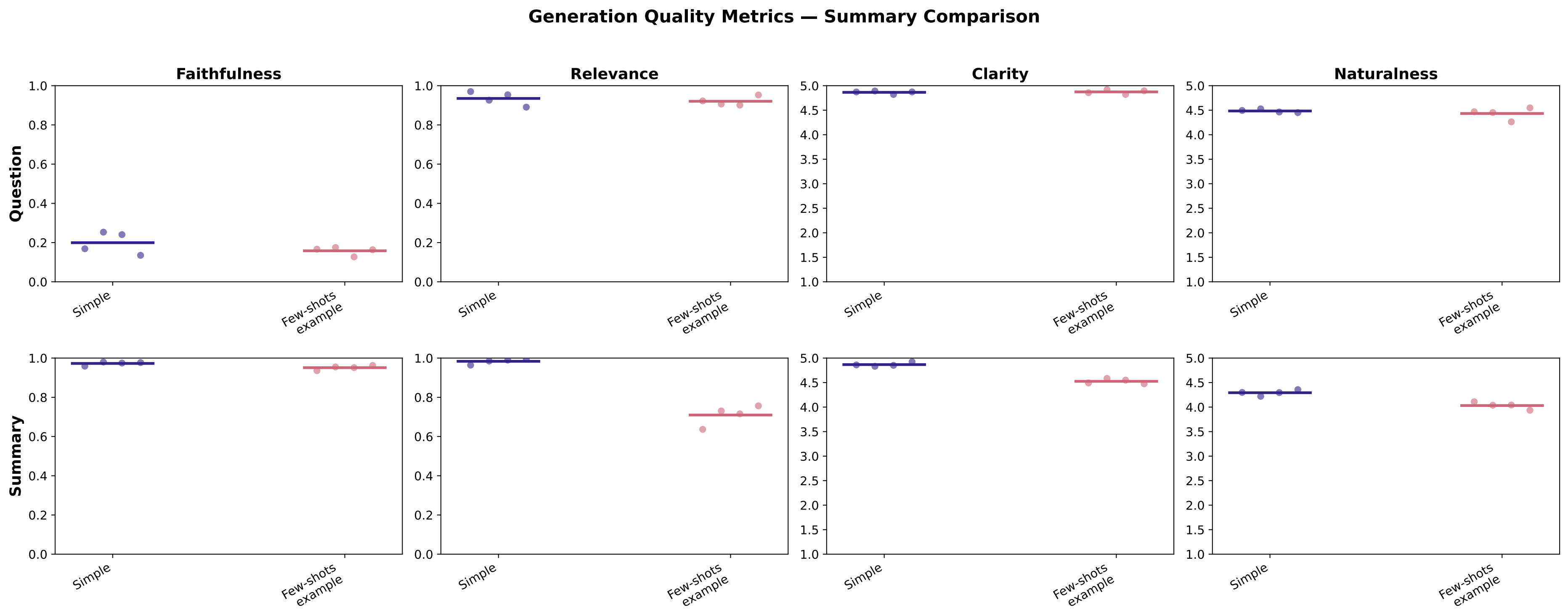


Supplementary Figure 15: **Comparison of the generative performance between a simple summarization prompt and a few-shot summarization prompt.** The enhanced summarization prompt does not influence the generation of the questions. However, with the few-shot prompt, the generated summaries tend to be less relevant, less clear, and less natural, reflecting the conciseness of the contact tracing notes. Both prompts' generation seems to exhibit similar faithfulness. Although the summaries seem to have a lower quality, the few-shot summary prompt seems to lead to better predictions (Supplementary Figure 16).

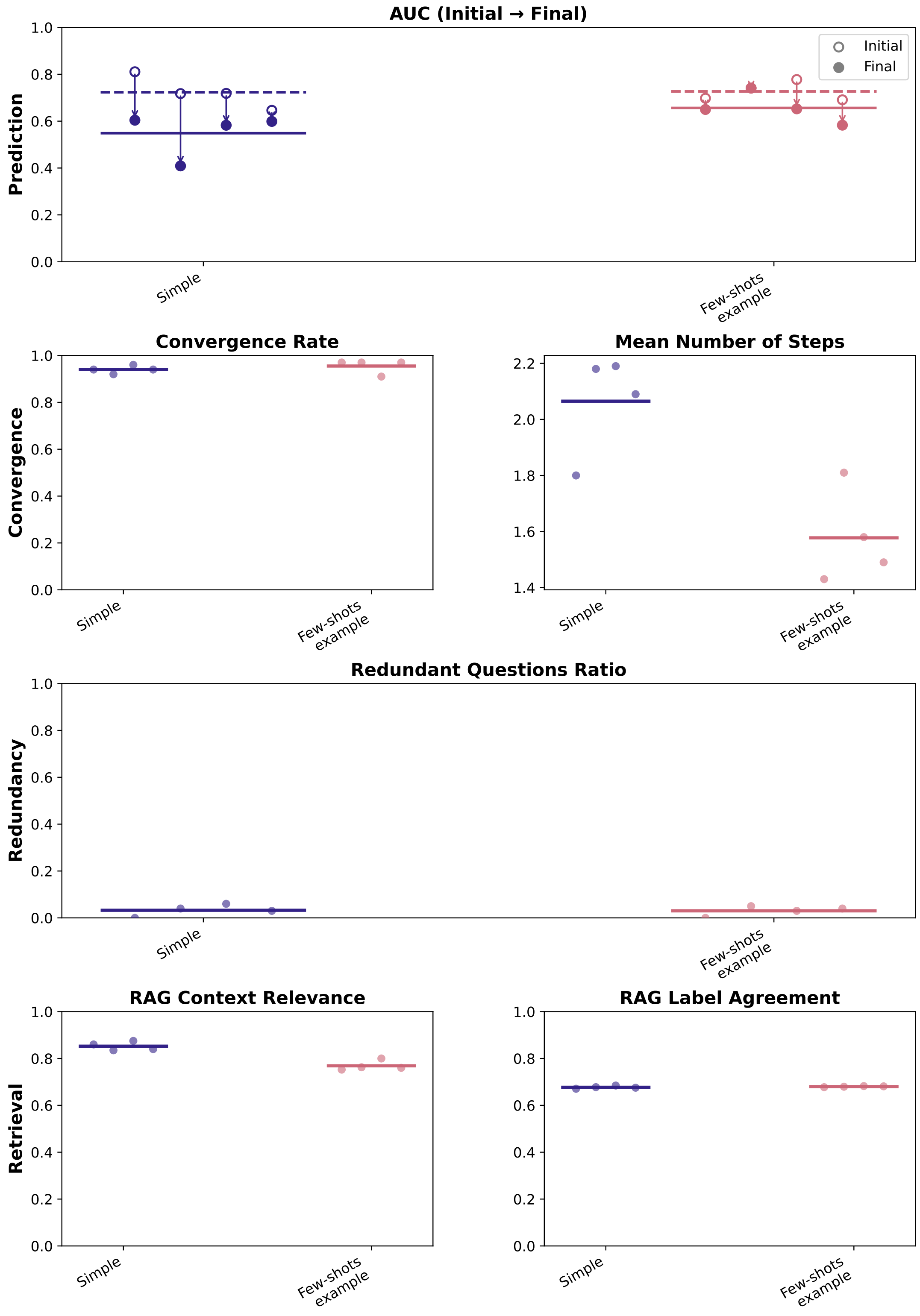


Supplementary Figure 16: **Comparison of the performance of the iterative agentic RAG framework at different steps between a simple summarization prompt and a few-shot summarization prompt.** Adding a few-shot example to the summarization prompt leads to better predictive performance over the iterations and converges in fewer steps. Our experiments with a few-shot example prompt did not show a strong influence on the questions' redundancy, nor on the retrieval performance.

### Appendix 13 Risk prediction discrimination across social contexts

We studied the behavior of the conversational framework with four different simulated synthetic profiles. Those represent distinct contact patterns: a low-contact profile (few social interactions), a high-contact profile (frequent and diverse interactions), a routine profile (recurring contacts in stable settings such as auditoriums and households), and a disrupted-routine profile (atypical contacts associated with travel). Among the $4 * 100$ conversations evaluated within our experiments, on average, the high-contact profile predicted a higher risk than the disrupted-routine profile, followed by the routine profile and the low-contact profile. The variance of the risk predictions increases with the average predictions among the synthetic profiles. Over the iterations with the higher-risk prediction profiles, we observe higher mean information gain. This gain is computed as $|P_k - P_{k-1}|$, where $P_i$ represents the predicted probability of infection computed after the $i^{th}$ interaction with the user. Finally, the convergence metrics differ between the different synthetic profiles. Indeed, the conversational agent converges slightly less often with the high-contact profile than with the other ones, which reflects more interactive steps on average with this profile. Interactions with the routine and disrupted-routine profiles involved a similar number of steps, which is higher than the average number of steps required for the low-contact profile before convergence (Supplementary Figure 17).

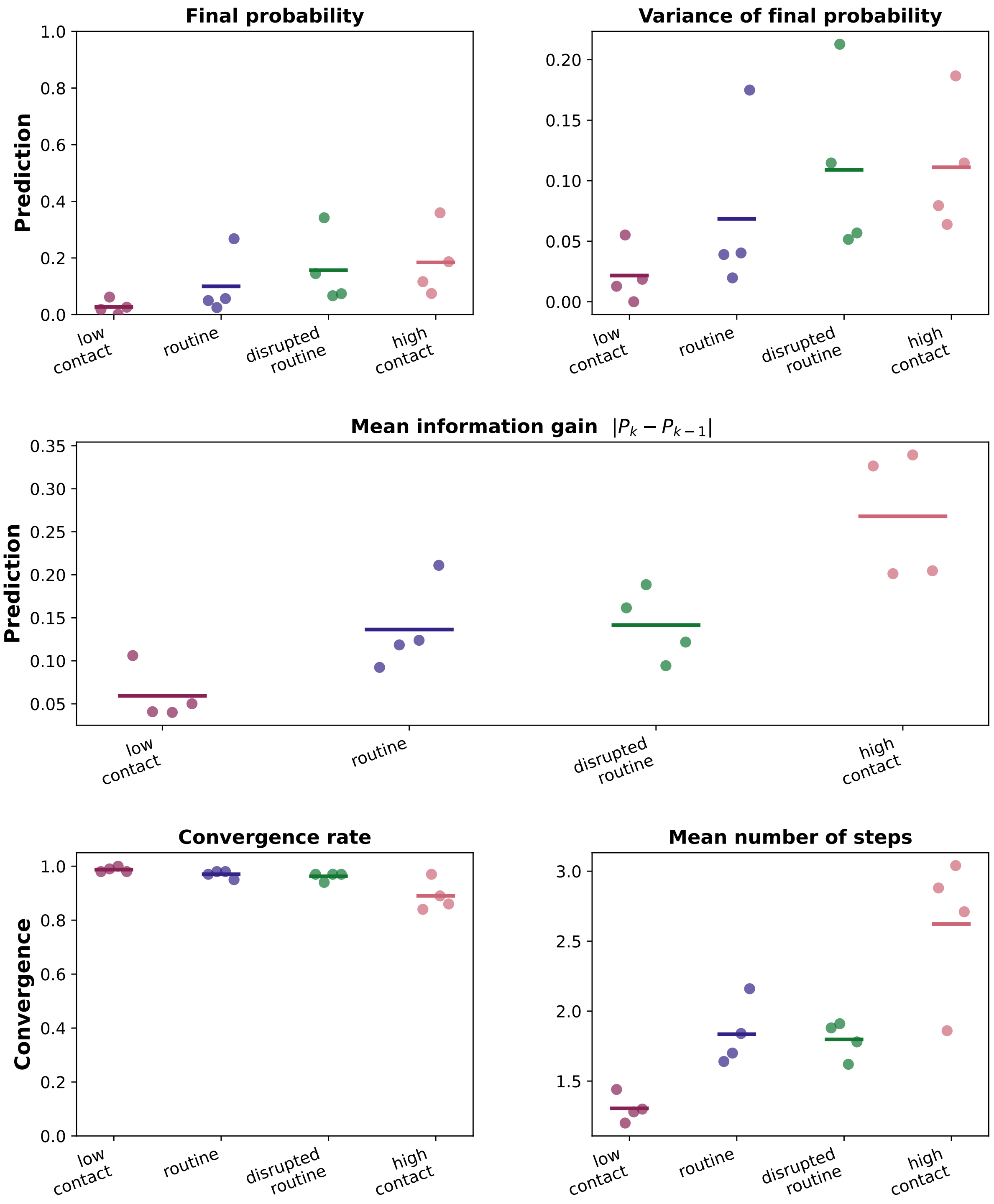


Supplementary Figure 17: **Comparison of predictive and convergence metrics between the four different simulated user types.** The final risk estimation seems to differ among the different user types. The higher the risk, the higher the variance among the risk predictions and the higher the mean information gain. Also, the predictions made for high-risk profiles required, on average, more iterative interactions with the conversational agent and converged less often.

As a secondary analysis, we also assigned a risk label ($\in \{0, 1, 2\}$) to each synthetic profile based on our own assessment. Based on the social background provided to each user type, we assigned a subjective risk label of 0 to the low-contact profile, a subjective risk label of 1 to the routine profile, and a subjective risk label of 2 to the disrupted-routine and high-contact profiles. We computed the Spearman ($\rho$) and

Kendall ($\tau$) rank-correlation coefficients between the $4 * 100$ final risk predictions for each user and their assigned subjective risk labels. The rank of the risk predictions slightly correlates with the rank of our own risk perceptions ($\rho = 0.28$, $p = 4 * 10^{-31}$; $\tau = 0.22$, $p = 4 * 10^{-30}$), in the context of the COVID-19 pandemic, within the student population in Leuven. The low p-values reflect a high number of data points considered to compute the correlation coefficients (Supplementary Figure 18). This last analysis, although more subjective than the previous ones, suggests an agreement between the risk we perceived and the risk estimated by the conversational framework.

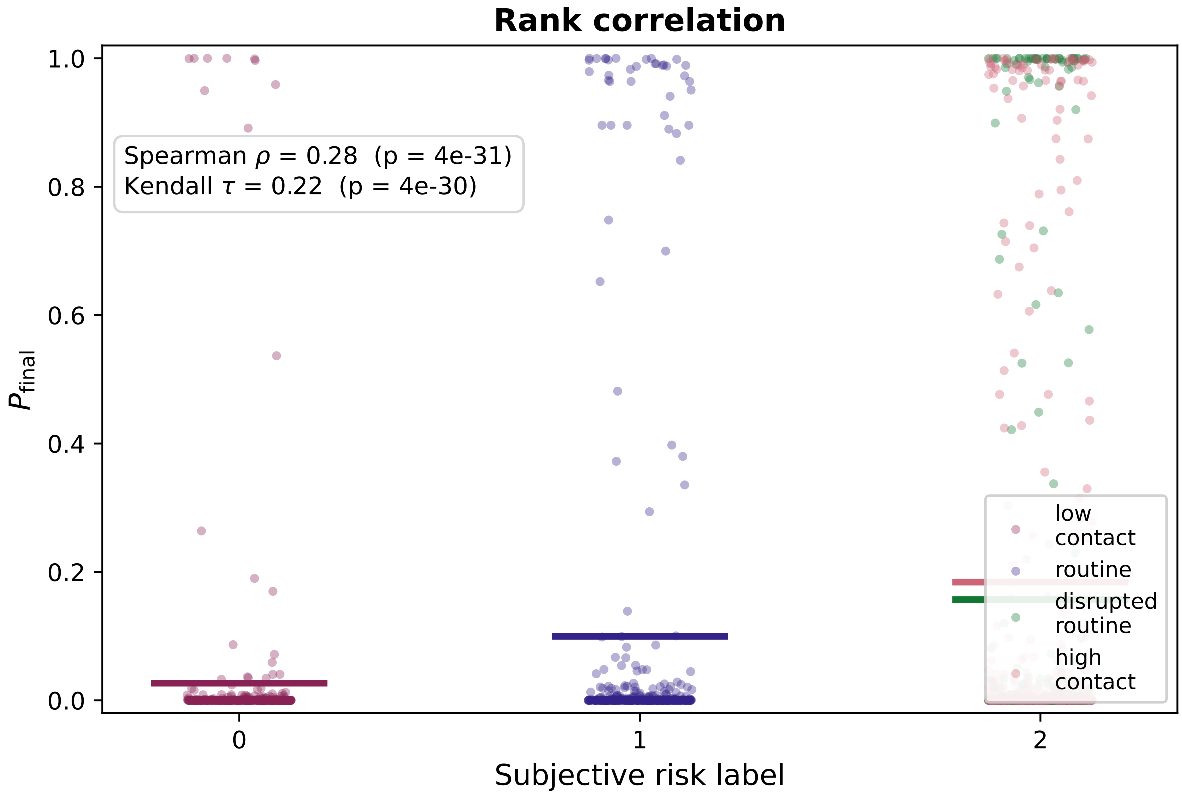


Supplementary Figure 18: **Rank-correlation between the risk of infection estimated by the conversational agent and the risk we perceived.** The subjective labels which we assigned to each user profiles slightly correlate with the final risk predictions computed by the conversational framework.

### Appendix 14 Example of a simulated conversation

This section presents a conversation between the main agent of our conversational framework and a simulated user. Both the agent and the simulated user were implemented with Llama3.3 (Supplementary Figure 19). Based on the initial description *"Scoutsweekend"* provided by the user, our framework retrieved words from the contact tracing dataset that may reflect a risk of infection in this social context (*"beerpong, openingscantus, dagelijkse, houseparty"*). The agent successively asks questions to gather information about those risk factors. The simulated user is prompted to answer "Ik weet het niet." if they do not know the answer based on the initial description of their social interactions. During the conversation, the framework refines the estimation of the risk of infection based on a generated summary of the conversation after each interaction. The framework identified a possible risk of infection when the simulated user mentioned singing during the activity (Supplementary Figure 20).

**Agent:** Beschrijf uw recente sociale interacties.

**Simulated User:** Scoutsweekend

**Agent:** Hebt u tijdens die sociale interactie beerpong gespeeld?

**Simulated User:** Dat weet ik niet, want er is onvoldoende informatie over de activiteiten die plaatsvonden tijdens de scoutsweekend.

**Agent:** Heeft u tijdens dit evenement gezongen of aan een openingscantus deelgenomen?

**Simulated User:** Ja, we hebben tijdens het scoutsweekend verschillende liedjes gezongen rond het kampvuur.

**Agent:** Heeft u tijdens deze activiteit elke dag contact gehad met dezelfde groep mensen?

**Simulated User:** Ja, want scoutsweekend wordt meestal doorgehouden met een vaste groep.

**Agent:** Heb je tijdens de scoutsweekend een huisfeest bezocht?

**Simulated User:** Ik weet het niet.

Supplementary Figure 19: **Simulated conversation from an agent and a user over a scouts activity.**

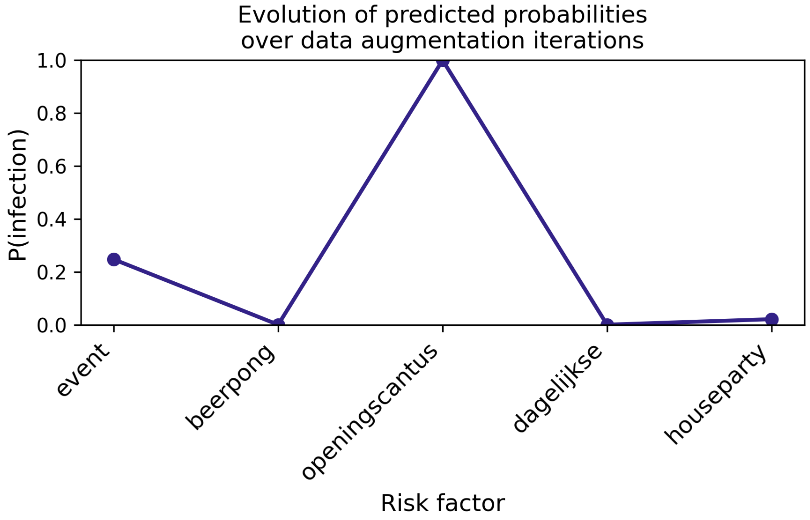


Supplementary Figure 20: **Evolution of the probability of infection estimated along the user's answers gathered with regard to the corresponding risk factors.** Those risk factors were used to generate questions in the conversation shown in Supplementary Figure 19.
